# Revealing geographic transmission pattern of COVID-19 using neighborhood-level simulation with human mobility data and SEIR model: A Case Study of South Carolina

**DOI:** 10.1101/2022.08.16.22278809

**Authors:** Huan Ning, Zhenlong Li, Shan Qiao, Chengbo Zeng, Jiajia Zhang, Bankole Olatosi, Xiaoming Li

## Abstract

Direct human physical contact accelerates COVID-19 transmission. Smartphone mobility data has been an emerging data source to reveal fine-grained human mobility, which can be used to estimate the intensity of physical contact surrounding different locations. Our study applied smartphone mobility data to simulate the second wave spreading of COVID-19 in January 2021 in three major metropolitan statistical areas (Columbia, Greenville, and Charleston) in South Carolina, United States. Based on the simulation, the number of historical county-level COVID-19 cases was allocated to neighborhoods (Census blockgroups) and points of interest (POIs), and the transmission rate of each allocated place was estimated. The result reveals that the COVID-19 infections during the study period mainly occurred in neighborhoods (86%), and the number is approximately proportional to the neighborhood’s population. Restaurants and elementary and secondary schools contributed more COVID-19 infections than other POI categories. The simulation results for the coastal tourism Charleston area show high transmission rates in POIs related to travel and leisure activities. The results suggest that the neighborhood-level infectious controlling measures are critical in reducing COVID-19 infections. We also found that the households of lower socioeconomic status may be an umbrella against infection due to fewer visits to places such as malls and restaurants associated with their low financial status. Control measures should be tailored to different geographic locations since transmission rates and infection counts of POI categories vary among metropolitan areas.

## 1 Introduction

A novel coronavirus was reported in late December 2019 in Wuhan, China (Nishiura et al. 2020), then was later identified as Severe Acute Respiratory Syndrome Coronavirus 2 (SARS-CoV-2). The coronavirus disease it caused was named COVID-19 (WHO, 2020). South Carolina (SC) in the United States (US) experienced four epidemic waves of COVID-19 from March 2020 to March 2022, causing 1.4 million cases and 17 thousand deaths. COVID-19 simulation and prediction, especially at the neighborhood level, plays an important role in health policymaking and disease prevention. COVID-19 simulation at the neighborhood level can identify the high-risk geographic locations with large numbers of new cases and help policymakers design appropriate disease control measures (e.g., mask wearing and vaccination policy) and social distancing policies tailored to these areas (Wrigley-Field et al., 2021). For instance, in adjunction with social distancing and mask wearing, prioritizing high-risk geographic neighborhoods for vaccination can effectively reduce COVID-19 transmission and mortality. Additionally, while the effectiveness of COVID-19 vaccine against the new variants of the virus is unclear, COVID-19 simulation and prediction could inform proactive personal protective measures to curb the disease transmission (Talic et al., 2021).

Direct human physical contact can accelerate COVID-19 transmission (Tian et al. 2020; Zeng et al. 2022; Zhang et al. 2020). Human mobility, as a proxy of human physical contact, shows a close relationship with disease spreading patterns (Hu T. et al., 2021) and thus has been used in COVID-19 simulation and prediction studies. For instance, Zeng et al. (2021a) predicted the 3-, 7-, and 14-day COVID-19 incidence at state- and county-levels in SC using Twitter-based mobility data. Subsequently, they also examined whether the impact of human mobility on COVID-19 incidence differed by communities with different proportions of older adults (Zeng, Zhang, Li, Sun, Yang, et al. 2021). Similar mobility-based investigations were applied based on various types of data sources at different geographic scopes. Hu S. et al. (2021) applied travel statistics from mobile devices (i.e., trip per person, person-miles traveled, and proportion of staying in homes) to model the effectiveness of non-pharmaceutical interventions in the US. Fritz et al. (2022) trained machine learning and statistical regression models using Facebook Social Connectedness Index (SCI) to predict COVID-19 cases in Germany. The historical mobility of Twitter users was also used to predict the worldwide spatiotemporal spreading of COVID-19 (Bisanzio et al., 2020; Li et al., 2020).

While many studies used mobility data to explore the patterns of COVID-19 transmission, most focused on relatively large scales such as countries (Hu T. et al., 2021), states (Zeng et al., 2021a), or counties (Zeng et al., 2021b). Only a few tried to simulate and predict the spreading using fine-grained (small geographic areas) mobility data. Among those, Chang et al. (2021a, 2021b) applied the sampled cellphone mobility data between neighborhoods and points of interest (POIs) and then derived transmission rates and infection counts for each neighborhood (i.e., census block groups) and POI of the top 10 largest metropolitan statistical areas (MSAs). The researchers essentially allocated the infections into neighborhoods and POIs using SEIR (*Susceptible, Exposed, Infection, Removed*) epidemiological models, which can identify the places of high transmission rates and incidence counts. These findings can inform evidence-based disease control measures tailored to the POIs with large numbers of infections. However, these two studies focused on POIs analysis only without reporting neighborhood transmission patterns. With the absence of the neighborhood level transmission patterns, the observation of POIs only may not reflect the overall spreading trend of COVID-19. In general, there is a dearth of studies exploring the relationships between human mobility and COVID-19 transmission at the neighborhood level, especially in less populous metropolitan statistical areas (MSAs). Their human mobility patterns may be different compared with the large ones such as New York MSA, which have been intensively studied (Chang, Pierson, et al. 2021; Yuan et al. 2022).

Research indicates that COVID-19 may have different transmission rates among population groups. For example, the infection rates of the elderly over 70 years were two times higher than the teenagers (10-19 years) (Davies et al. 2020); a higher proportion of Black race would increase the spread of COVID-19 (Zhai et al. 2021). A study on the early COVID-19 spreading in the northeast of the US found that counties with higher poverty and disability had lower rates of infection but higher death rates; the reason might be their lower mobility and higher comorbidities (Abedi et al. 2021). However, (Verma, Yabe, and Ukkusuri 2021) reported a contradictory observation in New York City and Chicago, where lower income groups had higher higher contact exposure and more cases. Levy et al. (2022) also documented the disparity of COVID-19 incidences among neighborhood socioeconomics, but they did not observe generality in the study area of three metro regions of the US (San Francisco, Seattle, and Wisconsin).

In this study, we aim to investigate the geographic transmission pattern of COVID-19 in three less populous MSAs in SC, which were under-studied in literature. The exploration on COVID-19 spreading in SC can fill the research gap for small MSAs. Neighborhood level (Census blockgroup, CBG) simulations of COVID-19 infections using human mobility data and SEIR epidemiological model were conducted in MSAs of Greenville-Anderson (Greenville), Charleston-North Charleston (Charleston), and Columbia. We estimated the transmission rates and infection counts for neighborhoods and POIs. Specifically, we aim to answer three questions:

1. Which neighborhoods and POI categories had high transmission rates of COVID-19? We are interested in which types of places where the residents or visitors are more prone to infect COVID-19 than others. Thus, high risk populations (e.g., the elderly) can stay away from those places in SC.
2. Which neighborhoods and POI categories have high COVID-19 incidence rates? Timely and appropriate responses can be applied to the places having high infection counts. For example, elementary schools with large infection cases may need special attention to decrease the spreading of the virus.
3. What are the correlations between the transmission rates and the socioeconomic status variables including demographic, social determinants of health, and visited POIs? Investigation of the associations of these variables could inform resource allocation and effective and precise disease prevention and control.

## 2 Methodology

This study used smartphone mobility data to explore the transmission patterns of COVID-19 in SC. The three most populous MSAs, i.e., Greenville, Columbia, and Charleston, were selected (Figure 1a). The study period is from December 29, 2020 to February 8, 2021, covering the main period of the second wave of COVID-19 spreading in SC (Figure 1b). We adopted a simulation model (Chang, Pierson, et al. 2021) based on mobility data and the SEIR model to estimate transmission rates and infection counts in neighborhoods and POIs. The evaluation (Section 2.3) for the simulation results contains three metrics: total case error, root mean square root (RMSE) of the daily case, and the error of infection rate by race (i.e., the White and Black). Based on the evaluated results, further analyses of the distribution of transmission rate and infection counts among CBGs and POIs were conducted (Section 2.4).

**Figure 1.**
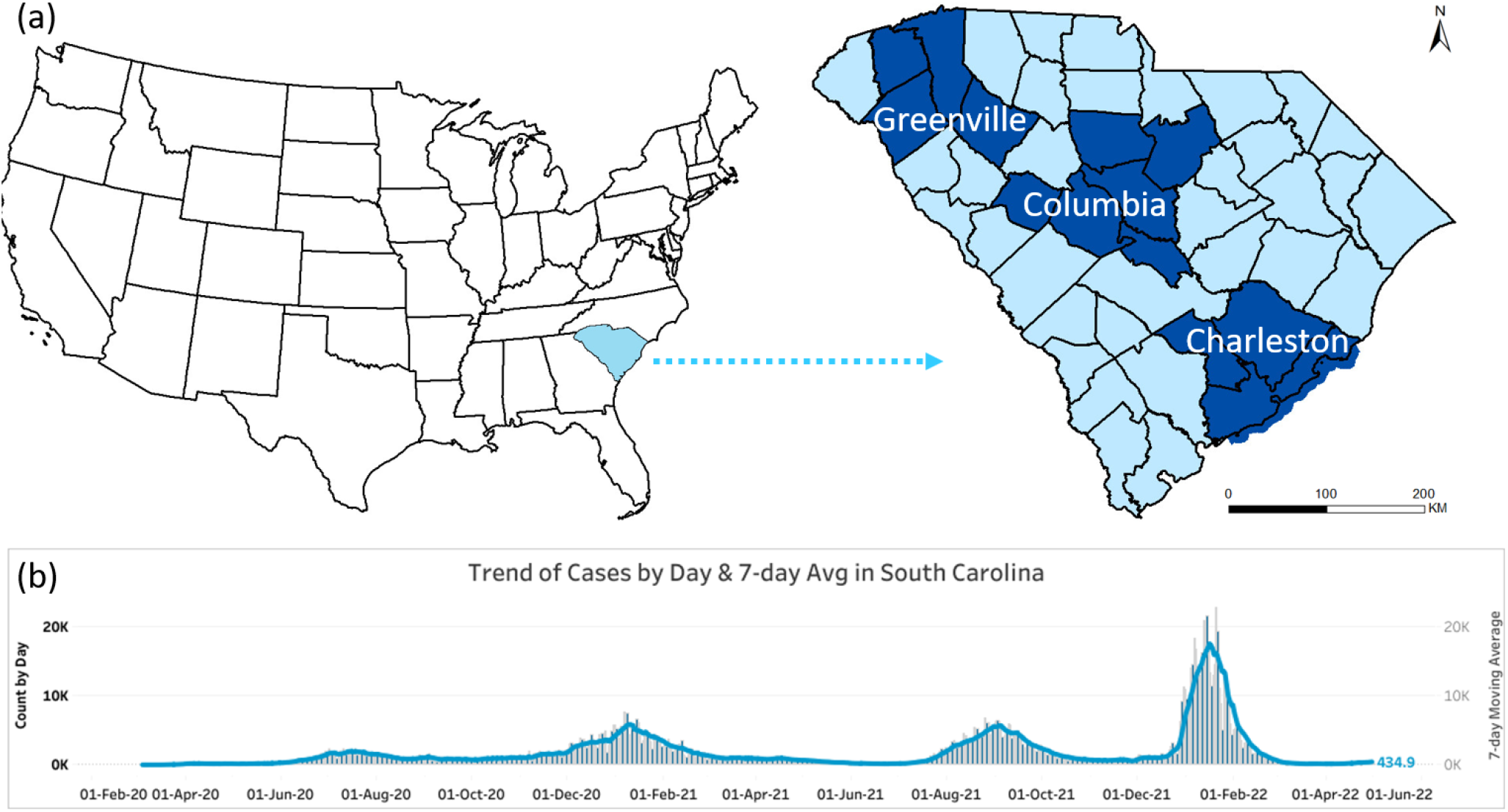
(a). Study area. The three selected MSAs are located in the northwest, middle, and southeast of SC; Charleston is a coastal area of developed tourism and shipping business. (b). Four COVID-19 waves in South Carolina as of June 2022. Credit: South Carolina Department of Health and Environmental Control (SCDHEC, 2022)

### 2.1 Datasets

SafeGraph datasets, including weekly *Patterns, Geometry*, and *Core Places* (SafeGraph 2022a). The Weekly *Patterns* contains the hourly visit records of POIs (e.g., restaurants, retail stores, and grocery stores) and the aggregated numbers of visitors’ home CBGs on that week. The Geometry dataset includes the areas of recorded POIs. The Core Places dataset has the basic information of POIs, such as the location name and category in the North American Industry Classification System (NAICS) (U.S. Census Bureau, 2022), including the top- and sub-category. About 10% of mobile devices in the US were sampled by SafeGraph (Squire 2019).

#### *New York Times historical COVID-19 data* (NYT COVID-19 data)

This dataset contains the daily cumulated confirmed COVID-19 cases at the county level in the US. It was obtained from github.com/nytimes/covid-19-data. The aggregated daily cases and total cases of MSAs were applied to evaluate the simulated results.

#### *American Community Survey 2019 5-year estimate* (ACS 2019)

It is the demographic estimation for each CBG. We applied the latest 5-year estimates when this study was conducted. Although ACS 2019 does not exactly align with the study period (January 2021), the population change is supposed to be too minor to overturn the simulation results.

#### *South Carolina County-Level COVID-19 Data by SCDHEC* (SCDHEC COVID-19 data)

This dataset (SCDHEC, 2022) provides county-level data of COVID019, containing test count, case count, and death count for age and race groups. The case counts of the White and Black race were used to assess those estimated from the simulation results.

### 2.2 Simulation model

We adopted Chang et al. (2021a)’s model to simulate the COVID-19 spreading in the study area. Necessary modifications were made to fit our study, such as neighborhood results storage because Chang et al. (2021a)’s work focused on POI only. The model assumes that people get infected merely in two types of places: their home neighborhoods (CBGs in this study) and POIs. The following introduces the simulation model.

Each CBG (say, ***c***_***i***_) has its own SEIR model, which maintains the number of individuals in 4 sequential stages for hour *t*: Susceptible 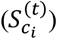, Exposed 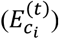, infectious 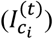, and Removed 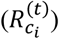; Figure 2 illustrates the evolution of 4 stages.

**Figure 2.**
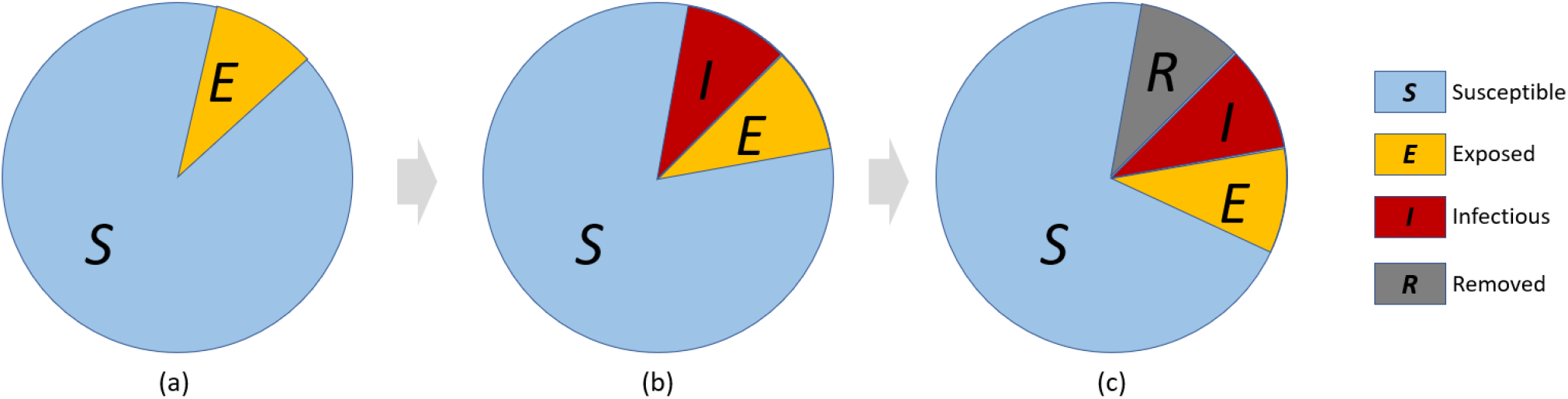
Evolution of 4 stages of SEIR model. (a): *Susceptible (S)* individuals have a probability (transmission rate) of being *Exposed* (*E*) to the virus; (b) the *Exposed* individuals become *Infectious* (*I*) after the latency period, and then make their close contacts become *Exposed*; (c) If the *Infectious* individuals in (a) are cured or dead, they become *Removed* (*R*) after the infectious period; *Exposed* individuals in (b) become newly *infectious*, while some *Susceptible* individuals become newly *Exposed*.

For CBG *c*_*i*_, its population 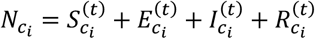, and the transition between four stages at hour *t* are:

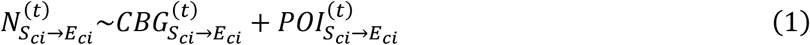

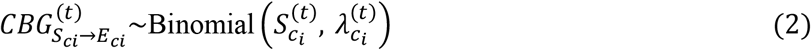

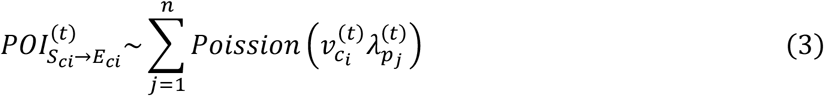

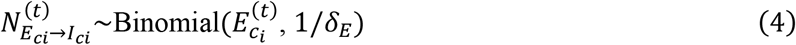

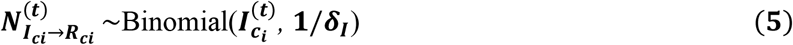

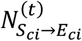 : the number of new exposures of CBG *c*_*i*_ at hour *t*.

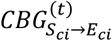 : new exposures occur in CBG *c*_*i*_ at hour *t*. Binomial 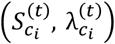 means the susceptible people 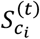 have a probability of 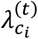 to be exposed.

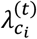: transmission rate of CBG *c*_*i*_ at hour *t*. 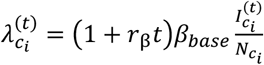, it is subject to a changeable coefficient (1 + *r*_*β*_*t*)*β*_*base*_ and the proportion of the infectious people in *c*_*i*_, i.e., 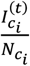. The denotation of β_*base*_ is the base transmission rate, shared with all CBGs; *r*_β_ is the slope of a linear function to capture the dynamic of β_*base*_. For example, if β_*base*_ does not change with *t, r*_β_ = 0; if β_*base*_ increases with *t, r*_β_ > 0, and vice versa.

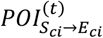 : new exposures of visitors from CBG *c*_*i*_ when they visit POIs at hour *t*. There are *n* POIs in total in a MSA.

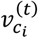 : the susceptible visitor number from CBG 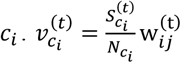, where 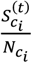 is the susceptible fraction of the population in CBG *c*_*i*_, 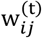 is the visitor count from *c*_*i*_ to POI *p*_*j*_ at hour *t*.

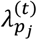: the transmission rate of POI *p*_*j*_ at hour *t*. The new exposures among those visitors follow distribution as *Binomial* 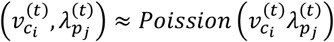, 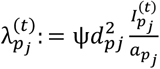, where *ψ* is the base transmission rate shared with all POIs, *d*_*pj*_ is the median dwelling time in POI 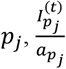 is the density of infectious visitors in POI 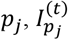 is the number of infectious visitors, and 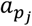 is the area of *p*_*j*_.

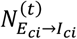 : the number of infected individuals in *c*_*i*_ at hour *t*. The model assumes that all *Exposed* individuals will become *Infectious*.

*δ*_*E*_: latency period.

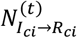 : the number of removed individuals in *c*_*i*_ at hour *t*. The model assumes that all *Infectious* individuals will become *Removed*.

***δ***_***I***_: infectious period.

In summary, there are three parameters in the simulation need to estimate: 1) *β*_*base*_, base transmission rate, shared with all CBGs; 2) *ψ*, the base transmission rate shared with all POIs; and 3) *r*_β_, the slope of the linear function to capture the dynamic of *β*_*base*_. The model used grid search to determine the optimal values of free parameters. We assumed that *r*_β_ would not change dramatically, so we set its range to [-0.5, 1]. Chang et al. (2021a) provide plausible ranges of *β*_*base*_ and *ψ* for the first COVID-19 wave of 10 metroplotian areas in the US impirically; however, these ranges did not fit the seconed wave SC in our experiments, so we adjusted them until the simulation generated results fitting to the reality. A grid search will use 1050 combinations of 10, 15, and 7 possible values with equal intervals for *β*_*base*_, *ψ* and *r*_β_, respectively. The parameter set whose simulation result is mostly close to the actual confirmed cases from NYT COVID-19 data is the best one, and it will be selected as the optimal model. Finally, we simulated each MSA respectively using the optimal model. A parameter table for the final model can be found in Table A1 in Appendix.

### 2.3 Evaluation metrics

The evaluation for the simulation contains three metrics: total case error, RMSE of the daily case, and the error of infection rate by race (i.e., the White and Black). The total case error is the ratio of the difference between the simulated case and NYT COVID-19 case to the latter. The simulated daily CBG cases were aggregated into MSA-level and compared with the NYT COVID-19 data to compute RMSE. The race infection rates were compared with SCDHEC COVID-19 data.

Equation (6) and (7) were used to compute the RMSE of the daily case. The simulated number of confirmed COVID-19 infections, i.e., 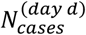, can be estimated by Equation (6) where *r*_*c*_ is the detected rate of infected individuals, *m* is the total number of CBG, and *δ*_*c*_ is the confirming lag, is set to 168 hours, or 7 days (Li et al. 2020). *D* in Equation (7) is the number of days in the simulation period, and 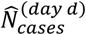 is the actual confirmed cases from NYT COVID-19 data. 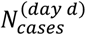 is the daily county-level aggregation of simulated infections in each CBG. The ground truth is 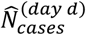,which was smoothed using a 14-days window to eliminate the confirmed cases fluctuation due to delay or correction. The simulation result of the smallest RMSE is preferred.

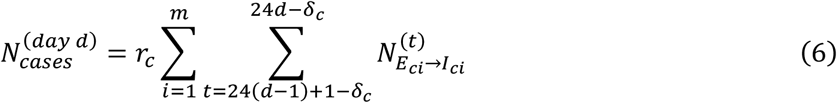

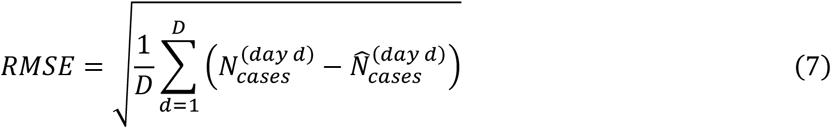

We ran 30 stochastic realizations of the binomial and Poisson distribution and then summed the outputs as the final number of the new 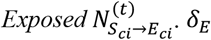 was set to 96 hours (Chang, Pierson, et al. 2021; Lauer et al. 2020), *δ*_*I*_ was 84 hours (CDC 2021; Li et al. 2020). *r*_*c*_ was empirically set to 65%. CDC (2021) has end estimation as 25% for *r*_*c*_ from February 2020 to September 2021 but this value contributed divergent results in our study period; we then tested a group of values and found that 65% can make the model convergent. The initialization of 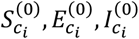, and 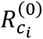 were according to NYT COVID-19 data: all *c*_*i*_ in the same county was assigned values according to their confirmed cases and population proportion to the county. Therefore, the 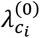 in the same county had the same value.

White and Black were the top 2 races in the study area, taking up 67% and 25% of the population. We estimated the White and Black cases using the product of simulated cases and race ratios of CBGs, then aggregated them into the MSA level. The race infection rate was the quotient of the race case divided by the race population. SCDHEC released COVID-19 cases by race but left about 20% of the total cases as *unknown*; we redistributed these *unknown* cases into race groups according to ratios of the known cases among races.

### 2.4 Transmission pattern analysis

After obtaining the optimal parameter set, its associated simulation results, including the hourly transmission rate and infection counts of each neighborhood and POI categories, were extracted for transmission pattern analysis. We computed the mean transmission rate and the sum of infection counts of each place and then compared CBGs and POI categories, respectively. The CBGs and POI categories having transmission rates and infection counts at the top ranks were identified as COVID-19 hot spots.

#### 2.4.1 Transmission rate analysis

This analysis focuses on the transmission rates of CBGs and POIs was applied to three MSAs, respectively. All hourly transmission rates, i.e., 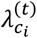 and 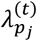 in the 30 stochastic realizations, were averaged as 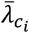 and 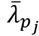, indicating the hourly mean COVID-19 transmission rates for CBG *c*_*i*_ and POI *p*_*j*_. Next, the mean transmission rate of each POI category, 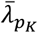, was calculated by averaging the 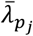 values of all POIs in this category. 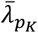 reflects the infecting risk when visiting the *K* category POI. In this study, the POI attribute of the top-category in NAICS was applied as the POI category (81 categories in this study). Finally, we mapped the distribution of 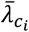 and compared the 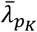.

#### 2.4.2 Infection count analysis

Infection count analysis investigated the distribution pattern of the number of infected individuals among CBGs and POI categories, respectively. The first step is to sum the hourly infection counts in the study period. Since the model assumes that all *Exposed* individuals will become *Infectious*, the hourly exposed counts of each place were summed up as the infection counts, denoted as 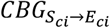 and 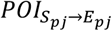. The former reflects the infection count that occurred in CBG *c*_*i*_, and the latter indicates the infection count occurred in POI *p*_*j*_. 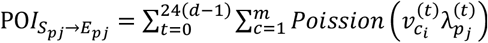. For each category, we have 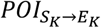 to present the sum of POIs infections in *K* category. According to Equation (1), for the residents in CBG *c*_*i*_, we can have 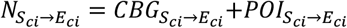, where 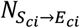 is the total infection count, and 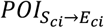 is the total infection count that occurred in POIs. Infection count analysis was also applied to the three MSAs respectively, and a comparison of the proportion of 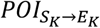 was conducted. Thus, we can observe whether MSAs have different patterns of infections via POIs.

#### 2.4.3 Correlation analysis

We conducted two Pearson’s correlation analyses to identify the association of socioeconomic status variables and COVID-19 spreading at the CBG level and POI level, respectively. At the CBG level, the correlation between the mean CBG transmission rate 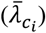 and the following variables were analyzed:

1. Demographic backgrounds, including population and proportions of the senior, White race, Black race, Hispanic race, and Asian race. The data source is ACS 2019.
2. Social Determinant of Health, containing the median household income, per capita building area, and proportions of poverty (population living below the federal poverty threshold), high-school diploma attainment (population of 25 years and over with the highest diploma from a high school), unemployed (unemployed civilian labor force), uninsured (population without health insurance), living with severe rent burden (household whose rent large than 30% of income), living with severe mortgage burden (household whose mortgage large than 30% of income).
3. The building area was computed using footprints generated from satellite images by a Microsoft research team (Microsoft, 2018); only residential buildings are kept according to OpenStreetMap land use data (OpenStreetMap 2022) and the SafeGraph Core POI dataset. The other variables came from ACS 2019.
4. POI characteristics, which consist of the mean transmission rate of visited POIs, per capita visit count to POIs, and infection count from POIs 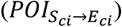, calculating from the SafeGraph datasets and simulated results.

At the POI level, we analyzed the correlation between the POI transmission rate 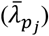 and the POI area, total visits, and infection count in POIs 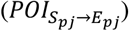. We removed the top 5% and bottom 5% values (outliers) of variables to keep the correlation analyses robust and ensure that the results reflect the correlation of the most values.

## 3 Results

### 3.1 Simulation and evaluation results

The number of cases from the simulation model successfully converged with the actual confirmed cases with small RMSEs. As illustrated in Figure 3, the simulated daily cases for each MSA (Greenville, Columbia, and Charleston) were close to the smoothed confirmed cases from NYT COVID-19 data; RMSEs were within 10% of the smoothed confirmed cases for all three MSAs (Table 1). The total simulated confirmed cases fit the actual cases within a minor error in the simulation period for all three MSAs: Greenville (−5%), Columbia (−2%), and Charleston (−7%) (Table 1). Both actual and simulated infections increased with the increase of the MSA population. During the study period, the number of POI visits decreased Sundays and dropped remarkably on Christmas day (December 25), but the weekday mobility trend is relatively stable (Appendix Figure A1).

**Table 1.** Simulation and evaluation results

|  | Greenville | Columbia | Charleston |
| --- | --- | --- | --- |
| **Population** | 895,942 | 824,278 | 774,508 |
| **CBG count** | 510 | 479 | 396 |
| **POI count** | 7,871 | 6,550 | 6,158 |
| **POI visit count** | 5,397,442 | 4,435,937 | 3,825,345 |
| **Confirmed cases** | 37,064 | 25,481 | 21,204 |
| **Simulated confirmed cases** | 35,166 | 24,760 | 19,702 |
| **Total case error** | -5% | -2% | -7% |
| **RMSE of daily case** | 67 | 54 | 47 |

Note. CBG: Census blockgroup; POI: Point of interest; RMSEs: Root mean square root.

**Figure 3.**
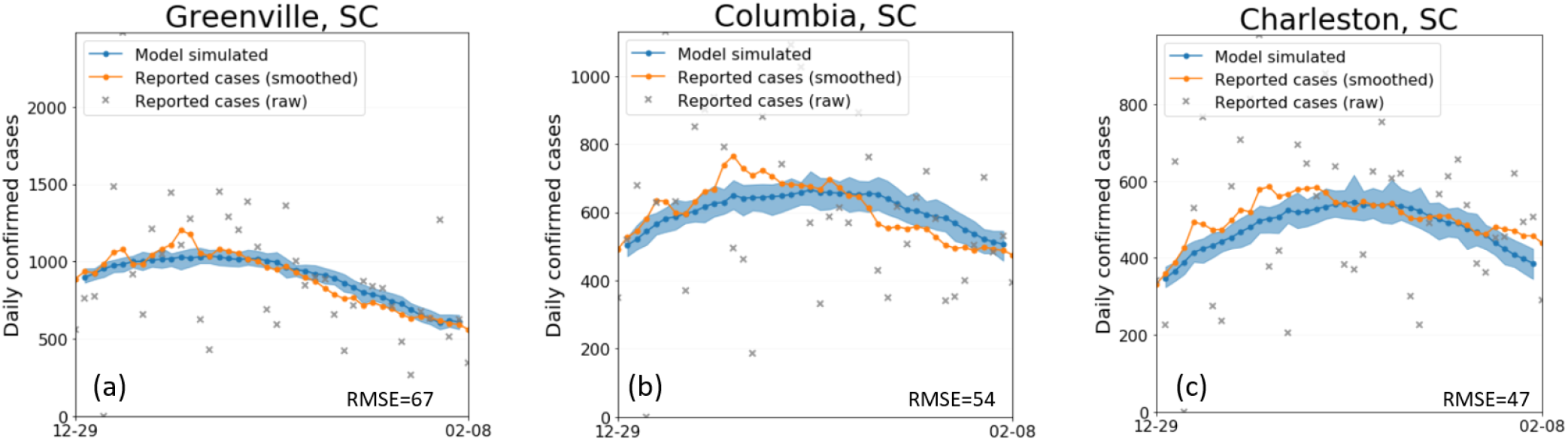
Simulation results of the daily cases and the RMSEs for the three MSAs. Shaded areas denote the 2.5^th^ and 97.5^th^ percentiles of the simulated daily confirmed cases from 30 stochastic realizations. SC: South Carolina; RMSEs: Root mean square roots; MSAs: Metropolitan statistical areas.

Table 2 shows the error of infection rate of the White and Black race between the SCDHEC COVID-19 data and simulated results. In the study period, the SCDHEC records show that the White and Black race have similar chances of getting infected (3.3% vs. 3.1%), and the simulated results accurately reflect this pattern, showing that the estimated race infection rates have minor gaps (0.1% – 0.4%) to SCDHEC records in each MSA.

**Table 2.**
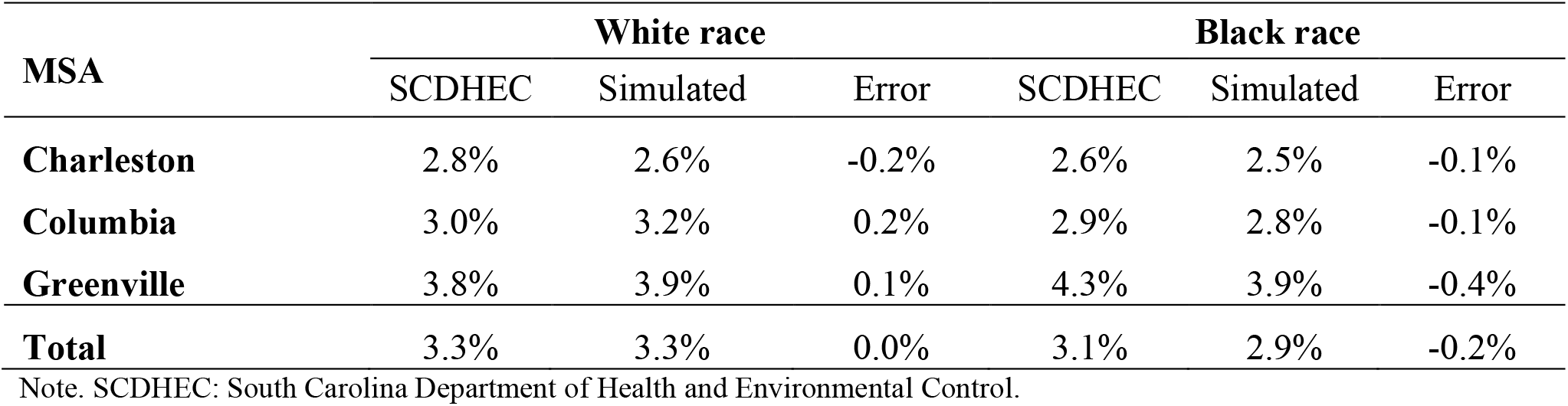
Infection rate error of the White and Black race

| MSA | White race |  |  | Black race |  |  |
| --- | --- | --- | --- | --- | --- | --- |
|  | SCDHEC | Simulated | Error | SCDHEC | Simulated | Error |
| **Charleston** | 2.8% | 2.6% | -0.2% | 2.6% | 2.5% | -0.1% |
| **Columbia** | 3.0% | 3.2% | 0.2% | 2.9% | 2.8% | -0.1% |
| **Greenville** | 3.8% | 3.9% | 0.1% | 4.3% | 3.9% | -0.4% |
| **Total** | 3.3% | 3.3% | 0.0% | 3.1% | 2.9% | -0.2% |

Note. SCDHEC: South Carolina Department of Health and Environmental Control.

### 3.2 Transmission rates distribution in CBGs

Figure 4 shows the mean transmission rates 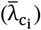 at the CBG level of the three MSAs. It reveals that the CBGs with high mean transmission rates are mostly scattered in Pickens County in Greenville MSA and Lexington County in Columbia MSA. The mean transmission rates show clear boundaries among countries due to the identical initialization inside a county. However, spatial patterns of the transmission rate within each county are also revealed as that some CBGs have higher or low transmission rates than their neighbors. For example, there is a clear hot spot in Pickens County (Greenville MSA) and a cold spot in Charleston County (Charleston MSA). Figure 4 reveals that residents face different infection risks among neighborhoods. Most CBGs in Greenville MSA had relatively high transmission rates, and Charleston MSA had low rates.

**Figure 4.**
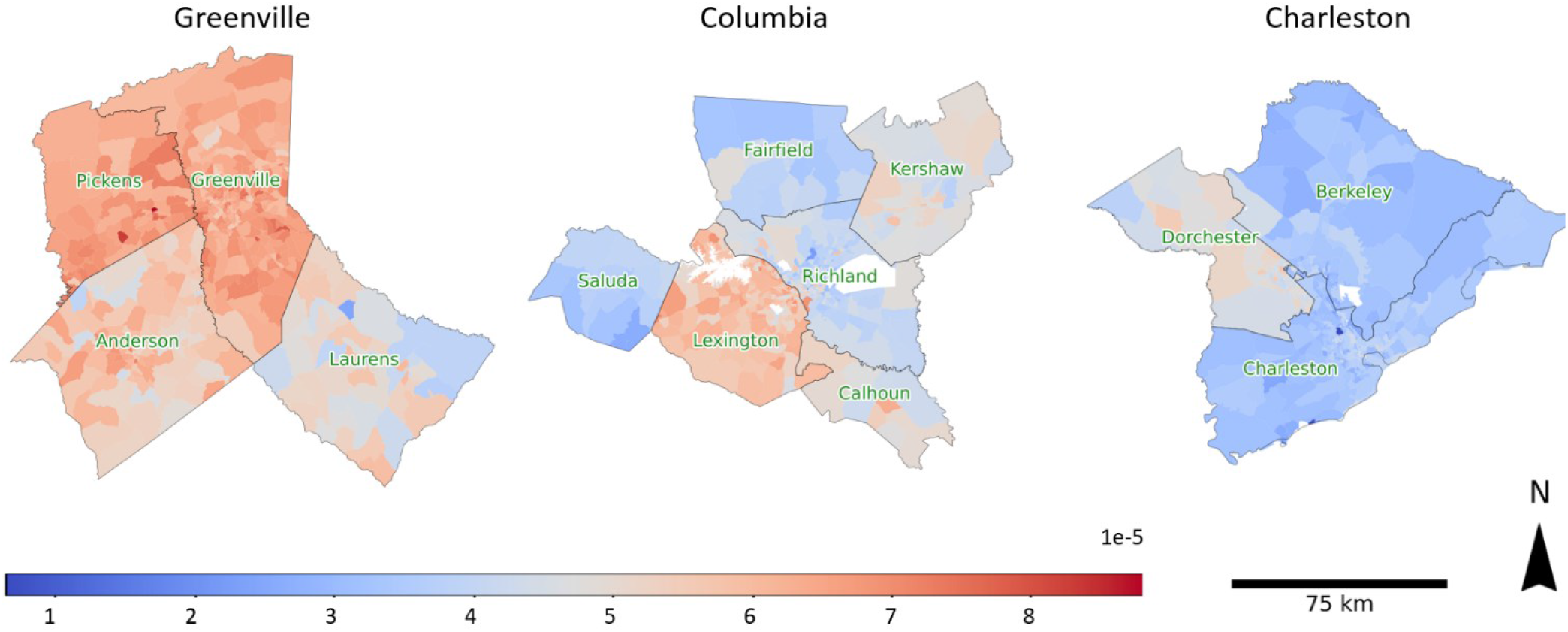
Mean transmission rates 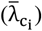 of CBGs. A high 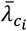 means a person has a high probability of infection in CBG *c*_*i*_ in an hour. (Blank areas are water bodies or military areas, which were excluded in this study). CBG: Census blockgroup.

### 3.3 Transmission rates distribution in POI categories

Figure 5 shows the mean transmission rates 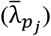 of the top 15 POI categories of the three MSAs. A high 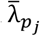 means a person has a high probability of being infected in POI *p*_*j*_ in a hour. The most infectious POI categories were *Support Activities for Road Transportation, Specialized Freight Trucking*, and *Home Health Care Services. Drinking Places (Alcoholic Beverages)* and *General Medical Surgical Hospitals* also had high transmission rates. Figure 6 further reveals that the transmission rate varies among MSAs. For example, POIs of *Support Activities for Road Transportation* in Charleston MSA had a remarkably higher transmission rate than the other two MSAs. In fact, most top 15 POI categories in Charleston have higher transmission rates. Interestingly, no category in the top 15 shares a similar transmission rate in the three MSAs.

**Figure 5.**
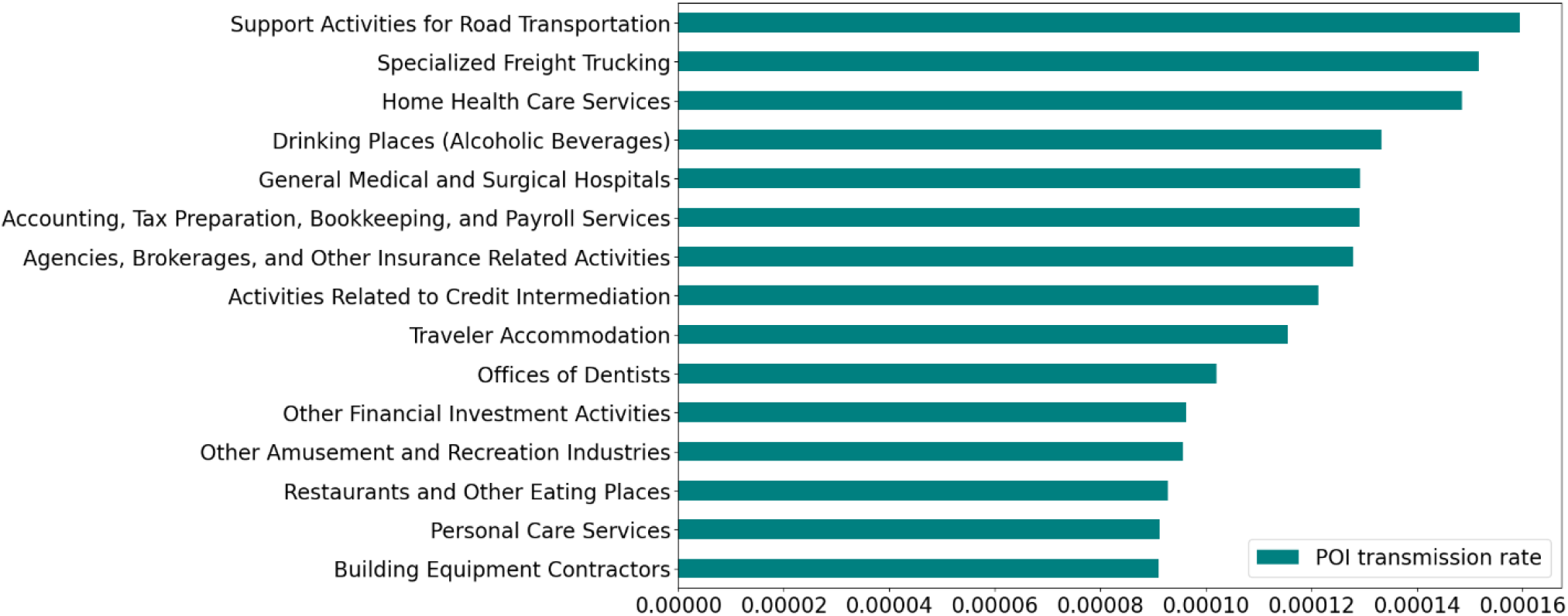
Mean transmission rates 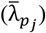 of top 15 POI categories. A high 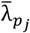 means a person has a high probability of infection in POI *p*_*j*_. POI: Point of interest.

**Figure 6.**
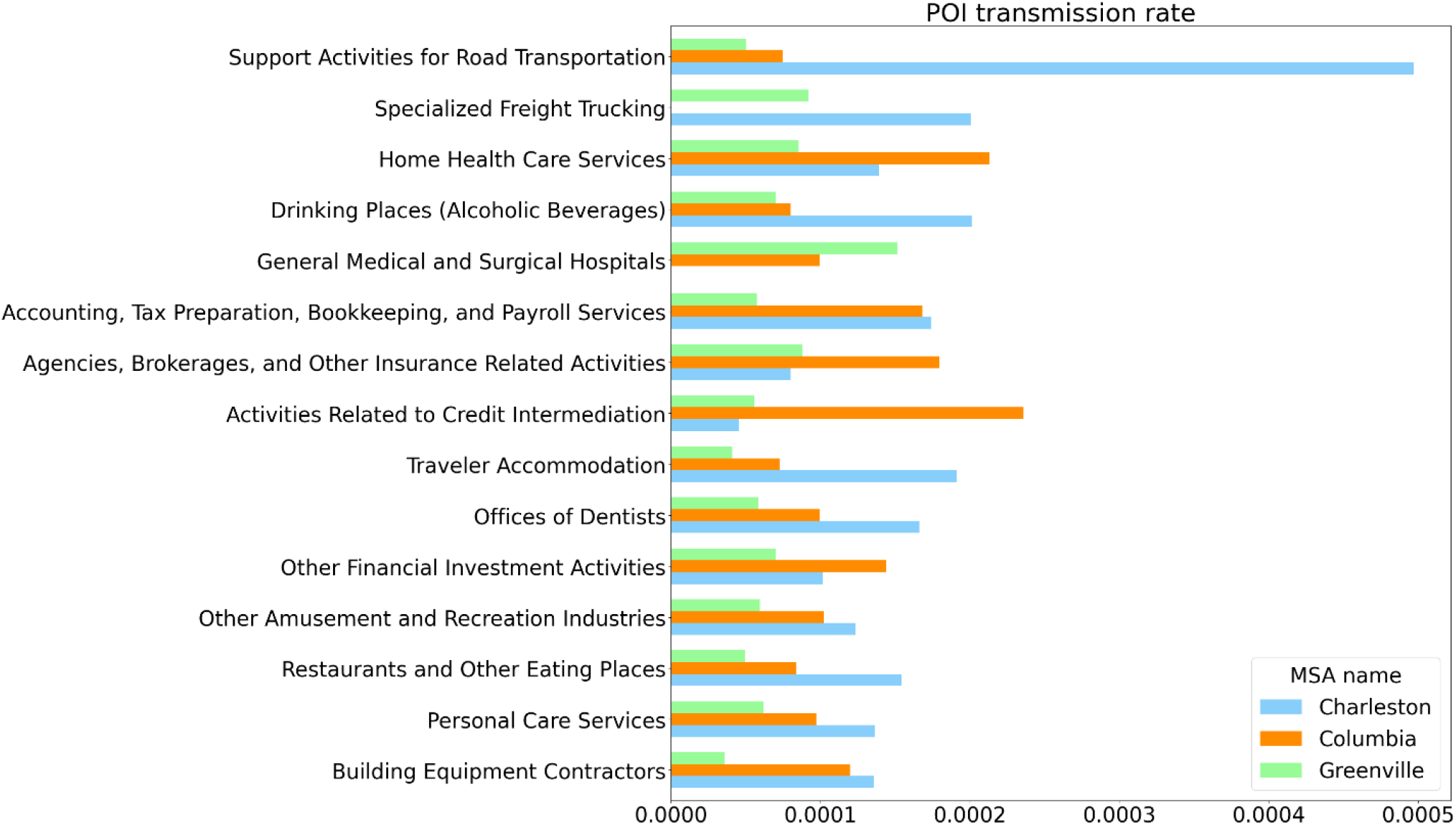
Transmission rates of top-15 POI categories among the three MSAs. POI: Point of interest; MSAs: Metropolitan statistical areas.

### 3.4 Infection counts distribution in CBGs

Similar to the CBG transmission rates, the simulated infection counts were not evenly distributed among CBGs (Figure 7). A few CBGs in central counties of MSAs had remarkably high infection counts; the peripheral counties or areas of MSAs had low infections. Averagely, a CBG had 88 COVID-19 infections in the study period. A few CBGs had dramatically higher infections than others, while most CBGs had low infections less than 100.

**Figure 7.**
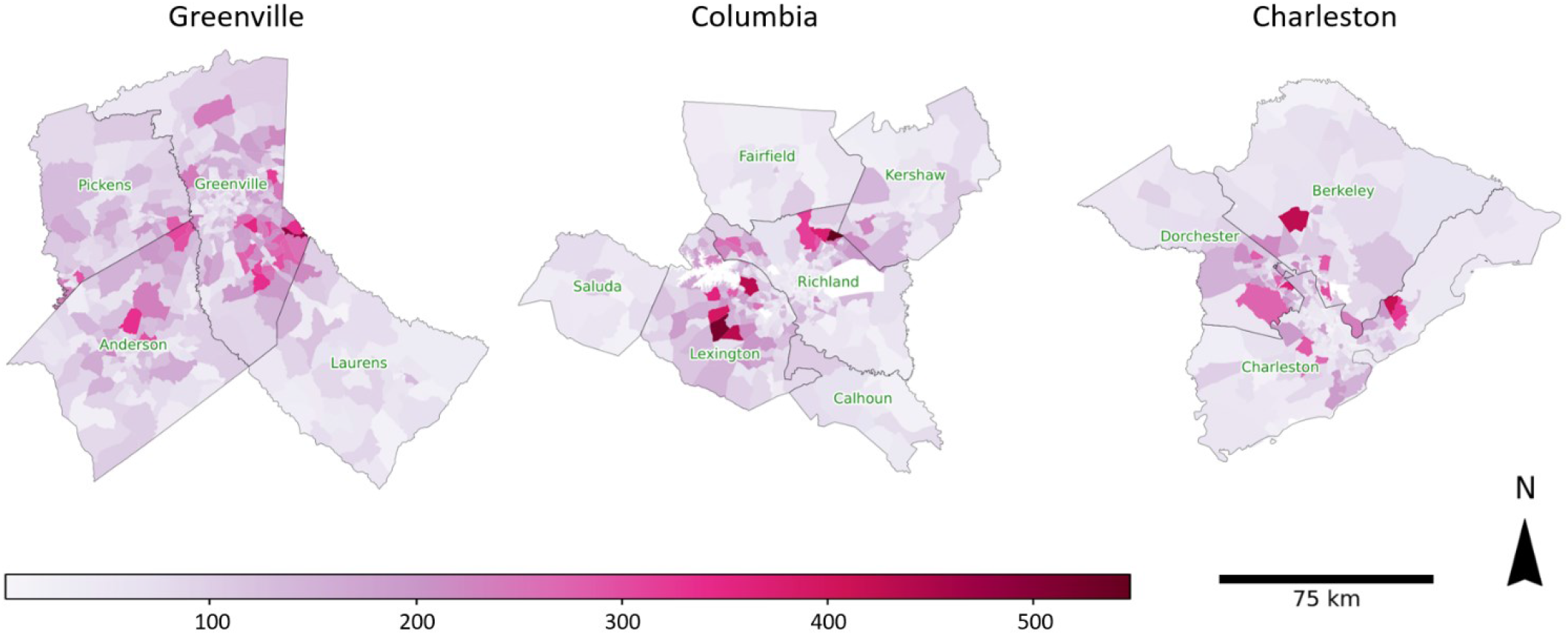
Infection counts distribution among CBGs

Table 3 shows the simulated infection counts that occurred in CBGs and POIs, i.e., the sum of 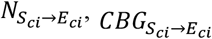, and 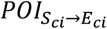 of each MSA in the study period. In the three MSAs, most simulated infections occurred in CBGs (86%). Charleston MSA had the highest POI infection count proportion (18%).

**Table 3.**
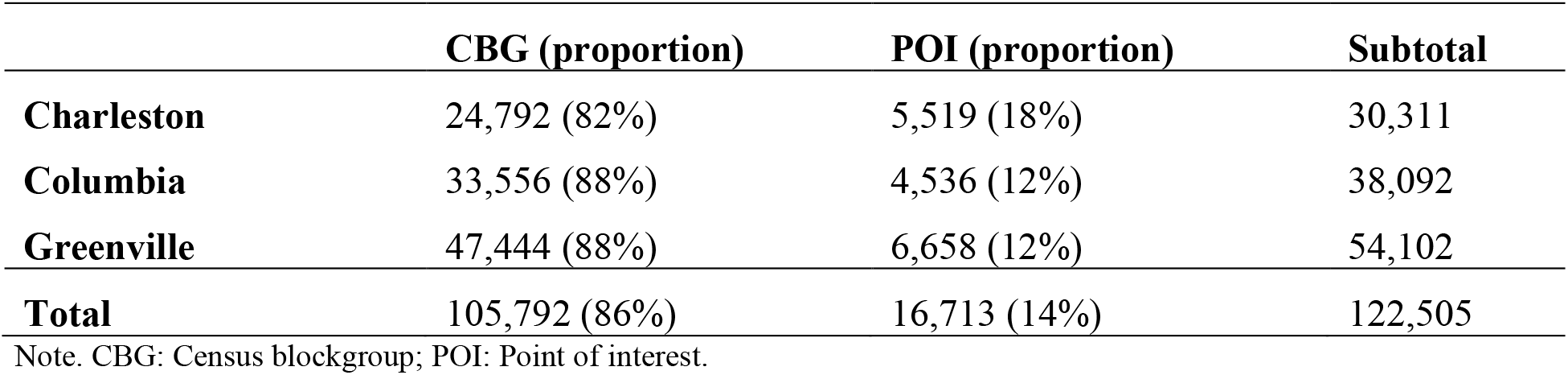
Simulated infection counts in CBGs and POIs

### 3.5 Infections in POI categories

Figure 8 shows the top 15 categories (i.e., *top-category* in NAICS) that occurred most COVID-19 infections in the simulation; most categories were the commonly visited categories, such as restaurants. It is worthy to note that the *Elementary and Secondary Schools* category ranked the second position, following the *Restaurants and Other Eating Places*; this trend was reflected by the official confirmed case (SCDHEC 2022): during the study period, COVID-19 infection in children and teenagers (<20 years old) was the highest share in SC (18.3%). *Other Amusement and Recreation Industries* and *Religious Organizations* ranked at the third and fourth place respectively.

**Figure 8.**
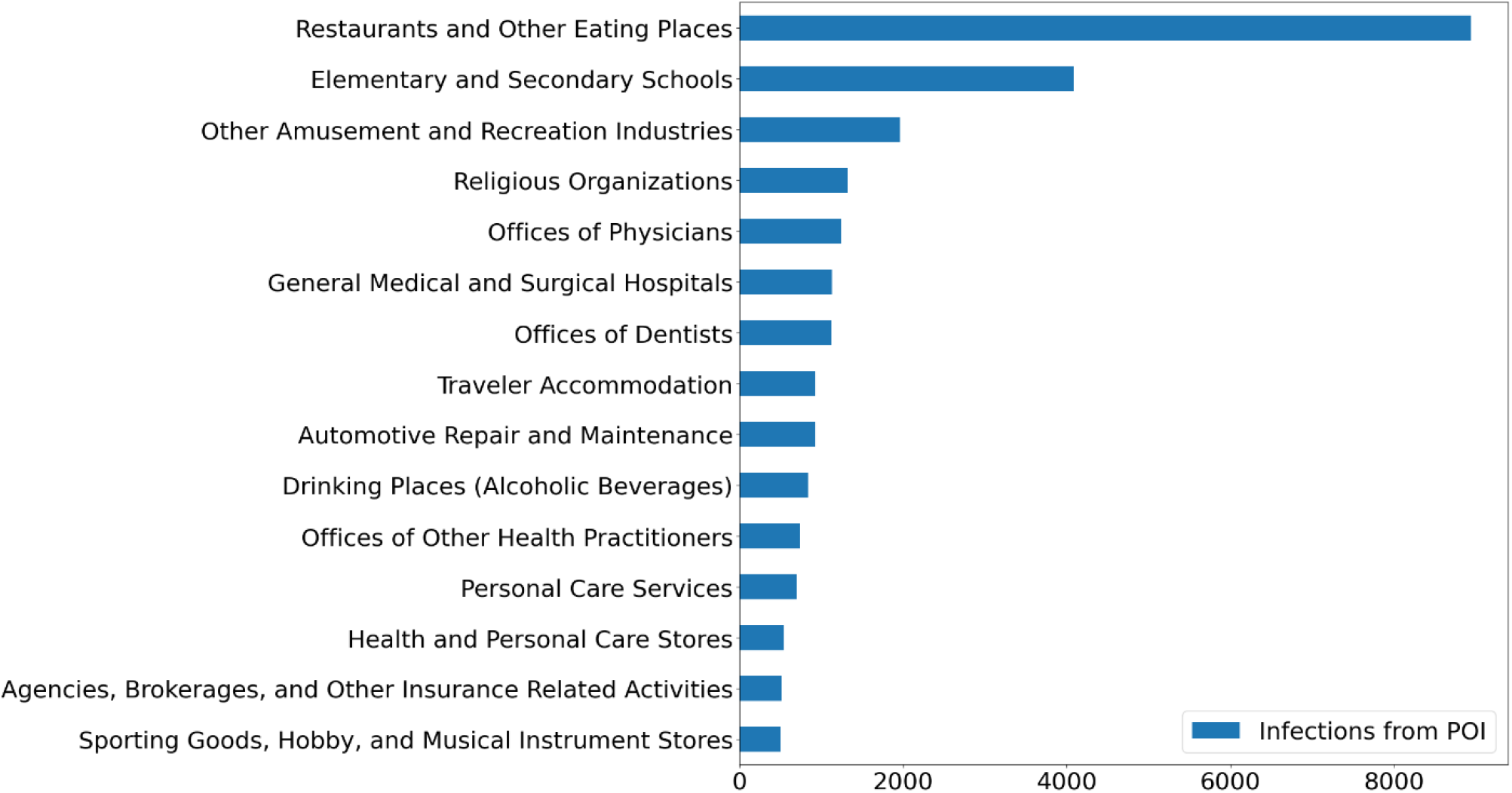
Infection counts from the top 15 POI categories of three MSAs.

When zooming into the MSA level, the infection counts show different patterns. For a better comparison between MSAs, POI category ratios were computed. The POI category ratio is the ratio of the infection count of a category to the total infection count of all categories in an MSA. Although *Restaurants and Other Eating Places* and *Elementary and Secondary Schools* still ranked in the first two positions, their category ratios vary among MSAs (Figure 9). For example, the category ratio of *Restaurants and Other Eating Places* was slightly more than *Elementary and Secondary Schools* (23% vs. 21%) in Greenville, but the difference between these two categories is much larger in Charleston (30% vs. 8%).

**Figure 9.**
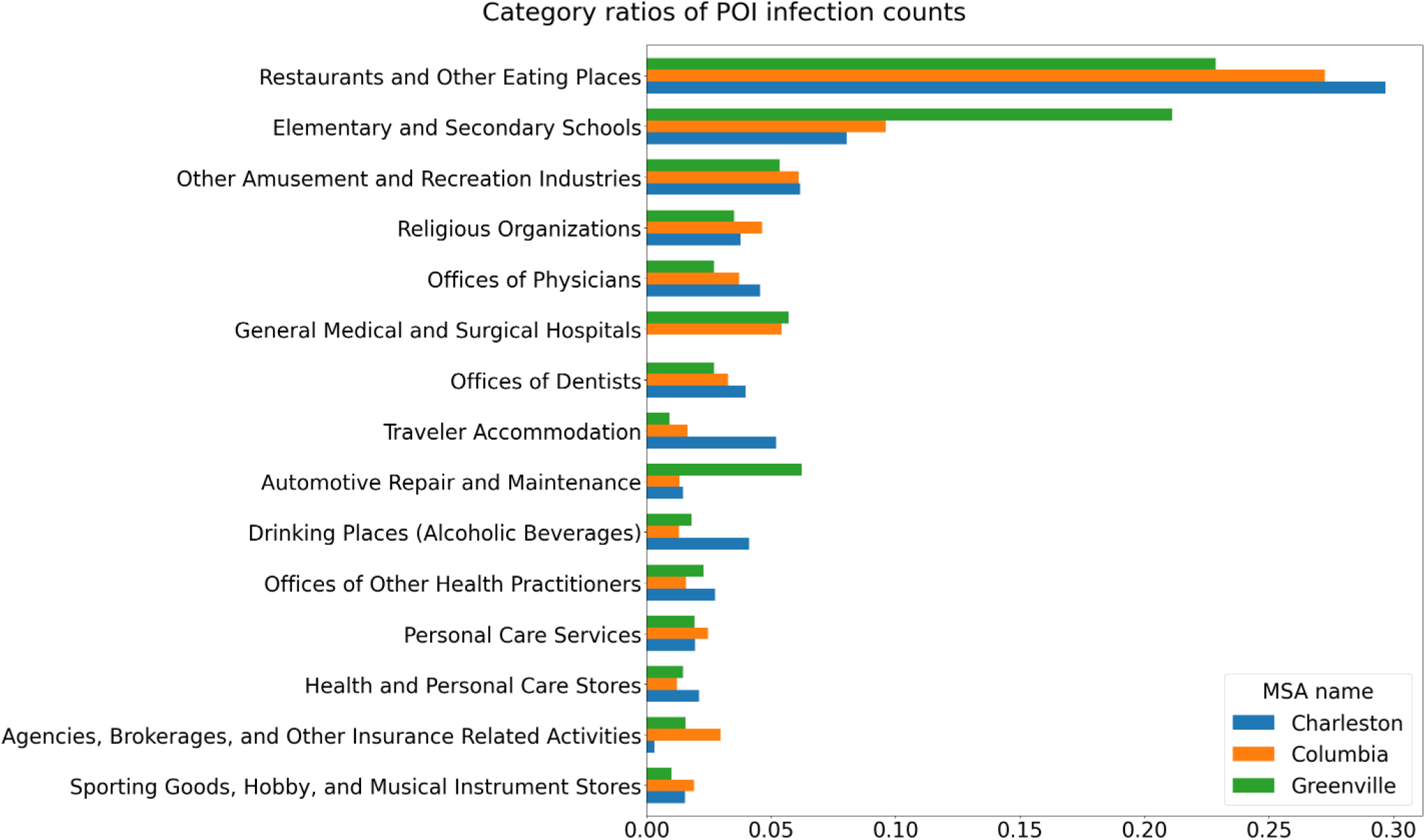
Infection ratios of the top 15 POI categories among three MSAs. (category ratio: the ratio of the infection count of a category to the total infection count of all categories). POIs: Points of interest; MSAs: Metropolitan statistical areas.

Similar to large MSAs in Chang’s simulation (2021a), the infection counts of POIs concentrated on top POI categories. For example, the top-5 categories across the tree MSAs took up 53% of infection counts, and the top 15% POIs consisted of 79% of infections.

Figure 10 shows the POIs with more than 30 infections that occurred in the MSAs. These POIs mostly located in the urban area of MSAs. This figure also reveals that POIs with high transmission rates did not necessarily have high infection counts and vice versa, although the mean POI transmission rate had a strong positive association with infections in POIs (see Table 5).

**Table 5.** Correlation analysis results of transmission rates at the POI level. The descriptive statistics of the variables are listed in Appendix Table A3.

| Variables | Pearson correlation coefficient ($r$) | | |
| --- | --- | --- | --- |
|  | Greenville | Columbia | Charleston |
| POI area (m<sup>2</sup>) | -0.137*** | -0.141*** | -0.167*** |
| Total POI visits | 0.127*** | 0.1268*** | 0.152*** |
| Infection count from POIs | 0.845*** | 0.847*** | 0.868*** |

Note: \* $p < 0.05$ , \*\* $p < 0.01$ , \*\*\* $p < 0.001$ ; POI: Point of interest

**Figure 10.**
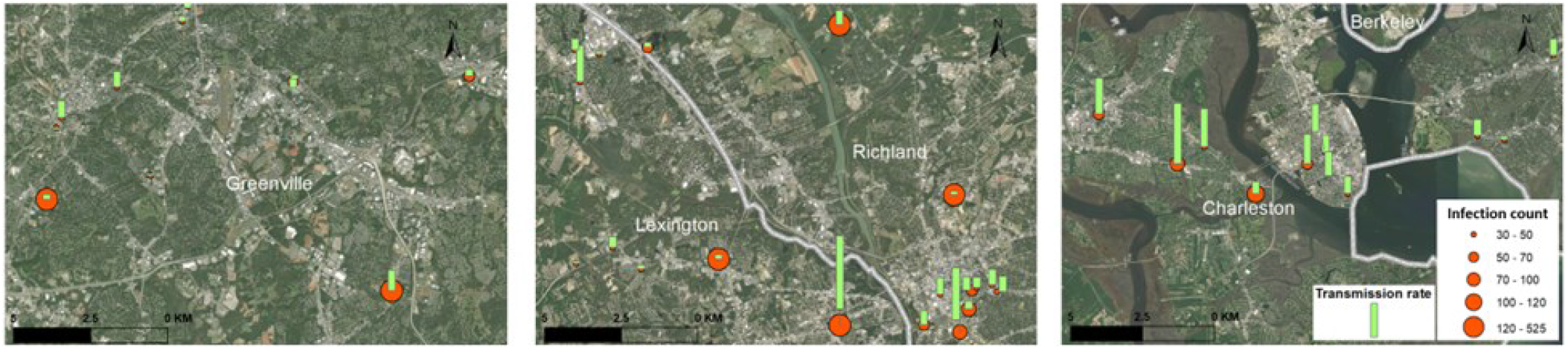
Spatial distribution of major POI spreaders in the three MSAs. POIs: Points of interest; MSAs: Metropolitan statistical areas.

### 3.6 Correlation analysis

Table 4 presents the correlation analysis results with the mean CBG transmission rates 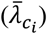. Several variables show significant correlations with CBG transmission rates, although the pattern varies among MSAs. For example, 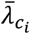 was significantly associated with population in Greenville (*r* = 0.264, *p* < 0.001) and Columbia (*r* = 0.133, *p* < 0.01) but not in Charleston. 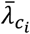 showed opposite associations with the mean transmission rate of visited POIs in Greenville (*r* = −0.124, *p* < 0.01) compared with Columbia (*r* = 0.429, *p* < 0.001) and Charleston (*r* = 0.403, *p* < 0.001). Per capita visit count to POIs had a positive correlation in Columbia (*r* = 0.425, *p* < 0.001) and Charleston (*r* = 0.174, *p* < 0.001), but not in Greenville. Per capita infection counts from POIs also have high positive correlations in Columbia (*r* = 0.442, *p* < 0.001) and Charleston (*r* = 0.344, *p* < 0.001) but demonstrated a weak negative correlation in Greenville (*r* = −0.091, *p* < 0.05).

**Table 4.**
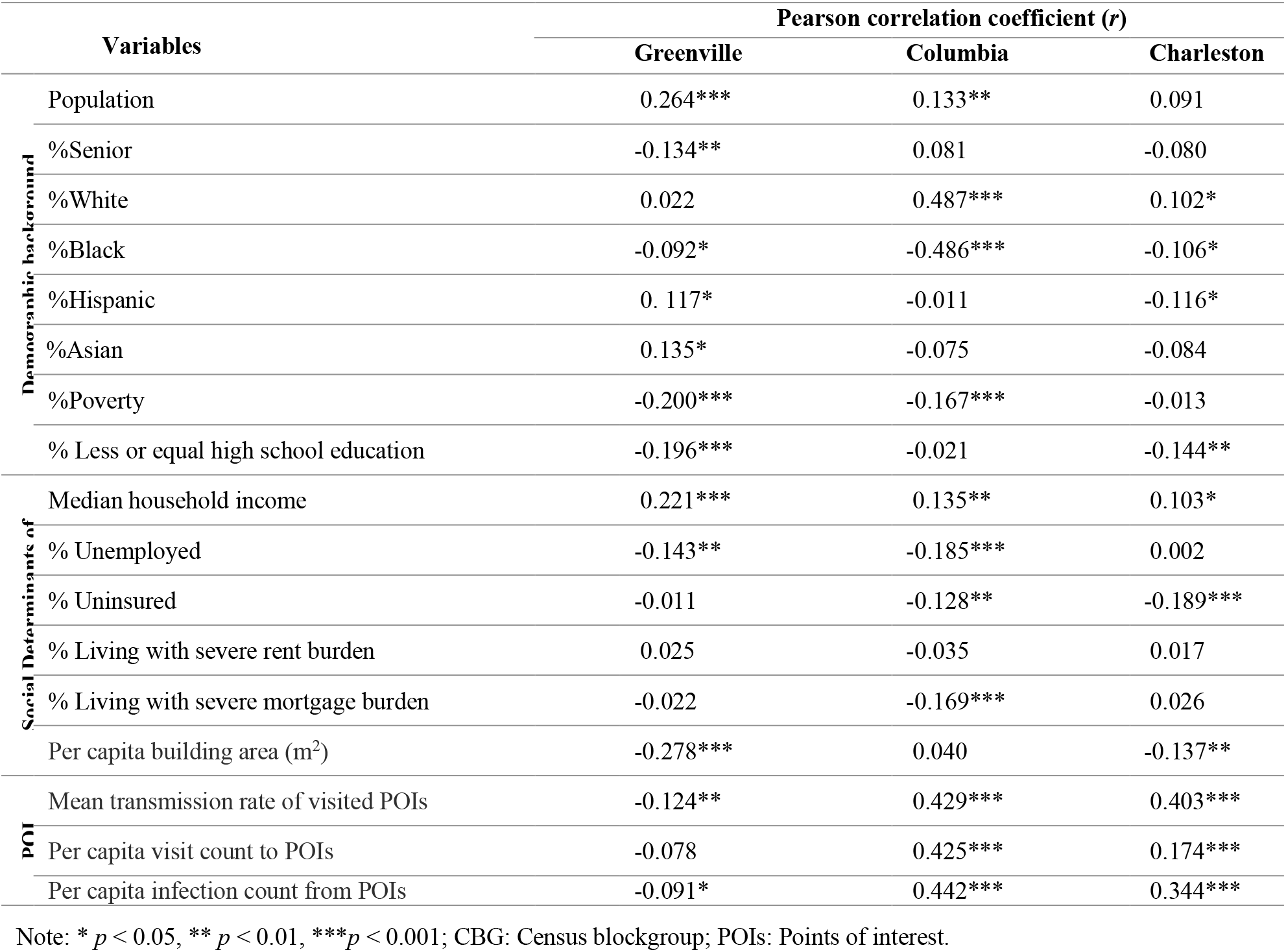
CBG level variables and their correlation with CBG transmission rates. The descriptive statistics of the variables are listed in Appendix Table A2.

In Columbia, 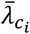 show much stronger positive correlation with the proportion of the White race (*r* = 0.487, *p* < 0.001) while Greenville and Charleston did not present significant associations. Since the White is the dominant race in quantity and the Black race was the largest minority in the study area, the correlation direction between 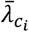 and the Black race proportion is the opposite of the White race. The median household income had a positive correlation with 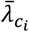 across all three MSAs (Greenville, *r* =0.221, *p* < 0.001; Columbia, *r* =0.135, *p* < 0.01; Charleston, *r* = 0.103, *p* < 0.05). The proportion of households living with severe mortgage burden tended to have a negative correlation with 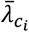 (*r* = −0.169, *p* < 0.001) in Columbia, but no significant association was found in Greenville and Charleston. The per capita building area shows negative correlations with 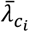 in Greenville (*r* = −0.278, *p* < 0.001) and Charleston (*r* = −0.137, *p* < 0.01). The scatter plots of CBG transmission rates and three selected variables (median household income, per capita visit count to POIs, and mean transmission rate of visited POIs) further revealed the varying correlation patterns among the three MSAs (Appendix Figure A5).

At the POI level, we analyzed the correlation between the POI transmission rate 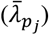 and POI area, total visits, and infection count in POIs 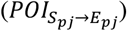. The result is shown in Table 5. The POI transmission rates had negative correlations with the POI areas and a positive correlation with visits to them. The infection counts from POIs had a strong correlation with POI transmission rates. All the three investigated MSAs shared similar patterns.

## 4 Discussion

### 4.1 The applicability of the mobility-based simulation method

The simulation of infectious disease transmission requires appropriate and robust human mobility data. In recent years, fine-grained SafeGraph mobility datasets have been one of the most used datasets for mobility studies. Our simulation in SC demonstrated that relatively small MSAs (ranking from 60 to 74 in population among 384 MSAs) can still benefit from a sparse mobility dataset. A dearth of studies has applied such datasets to small MSAs. For example, Chang et al. (2021a) focused on the top-10 MSAs in the US, which are more than eight times larger than the three MSAs in this study in the counts of CBGs and POIs. This study filled this gap, demonstrating the feasibility of using mobility-based simulation methods on small MSAs with low total case errors and low RMSEs of daily cases. The infection rates of the White and Black also matched the records of SCDHEC (ground truth), showing the reliability of the simulation results. The results of the validation analysis serve as the basis for the analyses of neighborhood level transmission rates, the number of cases, and associations.

### 4.2 Insights into measures against COVID-19 spreading based on the simulation results

#### 4.2.1 Effective control measures are needed to decrease disease transmission in neighborhoods

According to the simulation results, the infections that occurred in CBGs account for 86% of total cases, indicating that the disease control measures implemented at the neighborhood level might prevent COVID-19 transmission effectively. In January 2021, the restaurant restrictions in SC were lifted (McMaster and Governor 2020), and schools were reopened (SC Department of Education 2020). The surge of COVID-19 infections at the neighborhood level suggests that effective disease control measures tailored to high-risk geographic locations are needed, such as reducing parties and family gatherings or highlighting the importance of personal protective measures and vaccination. In addition, the concentration of infection counts and the strong positive correlation between infection count and neighborhood populations suggest that it is important to timely impose the control measures on the identified hot spots. Other less populous neighborhoods can have relatively easing measures to reduce the entire impact on ordinary life.

#### 4.2.2 The need for region-specific control measures

The simulation results reveal distinct patterns of COVID-19 transmission among the three MSAs in the second wave, suggesting that disease control measures tailored to different geographic locations need to be carefully developed by local authorities. For example, Charleston MSA is a popular coastal destination for US tourists, and its commercial shipping also plays an important role in local economy. The simulation shows that, compared with the other two MSAs, COVID-19 transmission rates in Charleston MSA are two times higher in the POI categories related to transportation and tourism, such as *Support Activities for Road Transportation, Specialized Freight Trucking, Drinking Places (Alcoholic Beverages), Traveler Accommodations*, and *Restaurants and Other Eating Places*. Therefore, the Charleston authority might need special restrictions on these POIs to balance the COVID-19 pandemic, disease control, and economic recovery. In Greenville, the ratios of infections in POI categories of *Elementary and Secondary Schools* and *Automotive Repair and Maintenance* were significantly higher than the other two MSAs. Appropriate measures are needed to reduce disease transmission via these two POI categories in Greenville MSA. Despite the noted difference, the ratios of infections in *Restaurants and Other Eating Places* among the three MSAs were similar (23%–30%) and ranked at the top position. Therefore, the restriction on restaurants remains the most universal and effective controlling measure regardless of region.

#### 4.2.3 Lower-socioeconomic status may act as an umbrella against COVID-19 in certain cases

The correlation analysis at the neighborhood level (CBG) shows that 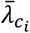 was positively associated with the lower socioeconomic status (SES) in the three MSAs, such as poverty, low rate of higher education, low median household income, unemployed, uninsured, and severe mortgage burden (Table 4), which is consistent with Abedi et al. (2021). CBGs with lower SES might have lower mobility (fewer POI visits) due to relatively low financial status, which results in a lower COVID-19 transmission rate. Mobility data from SafeGraph shows that the per capita visits to *Malls* and *Full-Service Restaurants* of the top decile CBGs in median household income were two times more than the bottom decile (Figure A3 in the Appendix). Similarly, the per capita time spent on *Nature Parks and Other Similar Institutions, Elementary and Secondary Schools* was also two times more than the bottom decile (Figure A4 in the Appendix). The observation of fewer POI visits in the low SES population at the CBG level in part explains why the White race has a higher infection rate than the Black since the White is associated with higher SES in the study area. For example, the percent of the White population in CBGs shows a positive correlation with median house income (*r* = 0.489, *p* < 0.001), and negative correlations with poverty (*r* = −0.437, *p* < 0.001), less higher education (*r* = −0.386, *p* < 0.001), unemployed (*r* = −0.355, *p* < 0.001), uninsured (*r* = −0.314, *p* < 0.001) and severe mortgage burden (*r* = −0.245, *p* < 0.001).

### 4.3 Implications in the disease control for future pandemics

The simulation results of the three MSAs in SC suggest that disease transmission modeling based on fine-grained mobility data can be used for forecasting and inform future disease control measures, offering a promising tool to support evidence-based public health emergency responses. First, the neighborhood- and POI-level simulation provides detailed spatial information on transmission rates and infection counts. High risk/vulnerable populations to COVID-19, such as senior residents, can avoid places of high transmission rates. Second, the local authorities can impose necessary disease control measures on neighborhoods and POIs which have large amounts of new cases to slow down the pandemic spreading. Third, the transmission patterns revealed in this study demonstrate that the transmission rate and infection count may vary significantly among MSAs; thus, there are no completely one-fits-for-all state-level disease control measures for different regions. Such simulation results can guide the local authorities to develop timely and appropriate measures tailored to different geographic and economic characteristics. Simulations based on fine-grained mobility data bring the opportunity to develop data-driven policy decision-making in developing and adapting emergency responses to pandemics and other public health emergencies.

### 4.4 Limitations and future research

While the findings are promising, two limitations of the study should be noted. The first limitation is the coverage of SafeGraph datasets. Though SafeGraph data has covered the entire US since 2018, it does not contain all POIs and is still being developed (SafeGraph 2022b). Another issue is data sampling. Although SafeGraph has a large proportion of mobile device – about 10% (Squire 2019), the sampling rate and reliability of visitation counts need further exploration. Further investigation is required to evaluate and calibrate the mobility matrix derived from SafeGraph data.

Another limitation is that the timing and initialization of the simulation may affect the results. Because the model assumed that there is a linear trend of the base transmission rate (*β*_*base*_) among all CBGs during the simulation period, the simulation period cannot be too long; otherwise, the linear trend may fail to capture the actual changing pattern of the base transmission rate. Therefore, our simulation cannot cover the entire second wave of COVID-19 in South Carolina; instead, we started at the time point (December 29, 2020) when daily cases have surged to 300 – 1000 in each MSA. Further research can introduce higher-order functions to present the dynamics of the transmission rate. Meanwhile, numerical optimization techniques are also needed to solve parameters of those high-order functions rather than grid search.

## 5 Conclusion

This study used fine-grained smartphone mobility data to simulate the COVID-19 spreading in the three MSAs of SC to obtain the transmission rates and infection counts at neighborhood- and POI-level. The aggregated confirmed cases in the simulation match the COVID-19 historical case trend at the MSA-level with low errors in three metrics: total cases, daily case RMSE, and the White and Black race cases, indicating that the simulating model can be used in relatively small MSAs which were under-investigated in the literature.

This study reveals that most simulated infections (86%) occurred in neighborhoods instead of POIs, suggesting that disease control measures in neighborhoods are critical to suppressing disease spreading during the study period. This imbalance of infection count between neighborhoods and POIs was rarely reported in previous studies. The patterns of transmission rates of neighborhoods and POIs significantly varied across MSAs; thus, general disease control measures might not be appropriate for each individual region. Our study shows that the simulation results can help local authorities develop effective and tailored measures according to regional geographic and economic characteristics. For example, the local authorities can advocate the high-risk and vulnerable population to avoid the places of high transmission rates and limit the human mobility surrounding certain neighborhoods and POIs with large numbers of new cases to curb the disease transmission. Therefore, the neighborhood-level simulations based on fine-grained mobility data and the SEIR model bring the opportunity to customize pandemic response in a data-driven manner.

## Data Availability

All data used in this study were retrieved from publicly accessible sources via the following links. SafeGraph mobility data: https://shop.safegraph.com/; New York Times historical COVID-19 data: https://github.com/nytimes/covid-19-data; American Community Survey 2019 5-year estimate: https://www.census.gov/data/developers/data-sets/acs-5year.html; South Carolina County-Level COVID-19 data by SCDHEC: https://scdhec.gov/covid19/covid-19-data/south-carolina-county-level-data-covid-19. The code for the study is provided at: https://github.com/GIBDUSC/covid-mobility-tool.

## Data availability statement

All data used in this study were retrieved from publicly accessible sources via the following links. *SafeGraph mobility data*: https://shop.safegraph.com/; *New York Times historical COVID-19 data*: https://github.com/nytimes/covid-19-data; *American Community Survey 2019 5-year estimate*: https://www.census.gov/data/developers/data-sets/acs-5year.html; *South Carolina County-Level COVID-19 data by SCDHEC*: https://scdhec.gov/covid19/covid-19-data/south-carolina-county-level-data-covid-19. The code for the study is provided at: https://github.com/GIBDUSC/covid-mobility-tool.

## Appendix

**Table A1:** Model parameters

| Parameter | Description | Value / Source |
| --- | --- | --- |
| $\beta_{base}$ | Base transmission rate among all CBG | Need to estimate |
| $r_\beta$ | Change rate of $\beta_{base}$ in the simulation period | Need to estimate in the range of $[-0.5, 1]$ |
| $\psi$ | Transmission scaling factor for all POI | Need to estimate |
| $\delta_E$ | Mean latency period | 96 hours (Chang, Pierson, et al. 2021; Lauer et al. 2020) |
| $\delta_I$ | Mean infectious period | 84 hours (CDC 2021; Li et al. 2020) |
| $\delta_c$ | Mean lag period to be confirmed | 7 days (Li et al. 2020) |
| $r_c$ | Detected rate of infection | 65%, empirically searched. |
| $N_{c_i}$ | Total population of CBG $c_i$ | ACS 2019 |
| $w_{ij}^{(t)}$ | Mobility matrix from CBG $c_i$ to POI $p_j$ at hour $t$ | SafeGraph |
| $a_{p_j}$ | Area of POI $p_j$ | SafeGraph |
| $d_{p_j}$ | Median visitor dwell time $p_j$ | SafeGraph |
| $S_{c_i}^{(0)}$ | Initial susceptible population in CBG $c_i$ | NYT COVID-19 data |
| $E_{c_i}^{(0)}$ | Initial exposed population in CBG $c_i$ | NYT COVID-19 data |
| $I_{c_i}^{(0)}$ | Initial infectious population in CBG $c_i$ | NYT COVID-19 data |
| $R_{c_i}^{(0)}$ | Initial removed population in CBG $c_i$ | NYT COVID-19 data |

**Figure A1.**
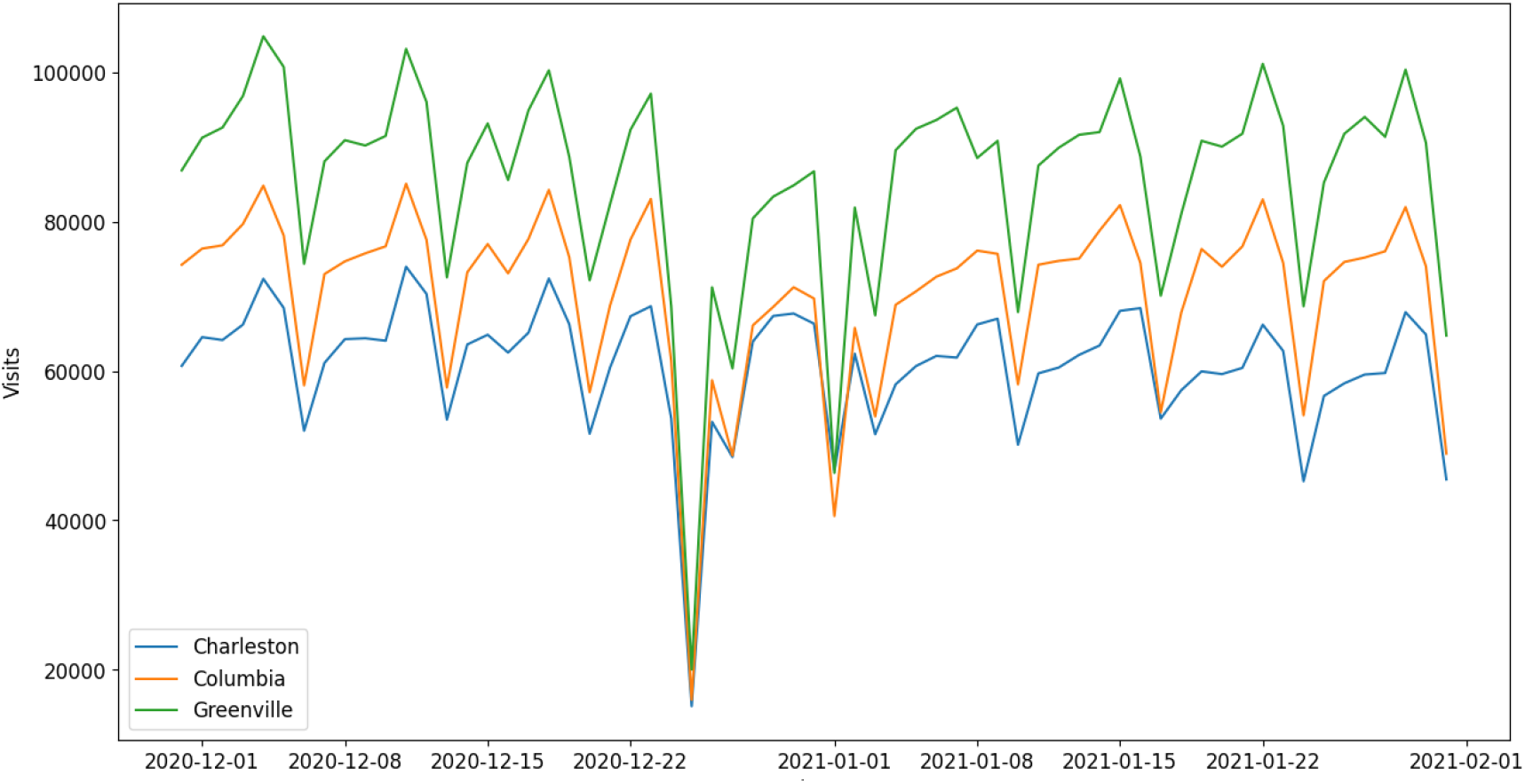
Number of visits was in a stable pattern during the study period. Daily POI visits have a weekly fluctuation pattern with a decrease on Sundays (troughs), and dropped remarkably on Christmas day (December 25, the lowest trough). POI: Point of interest.

**Figure A2.**
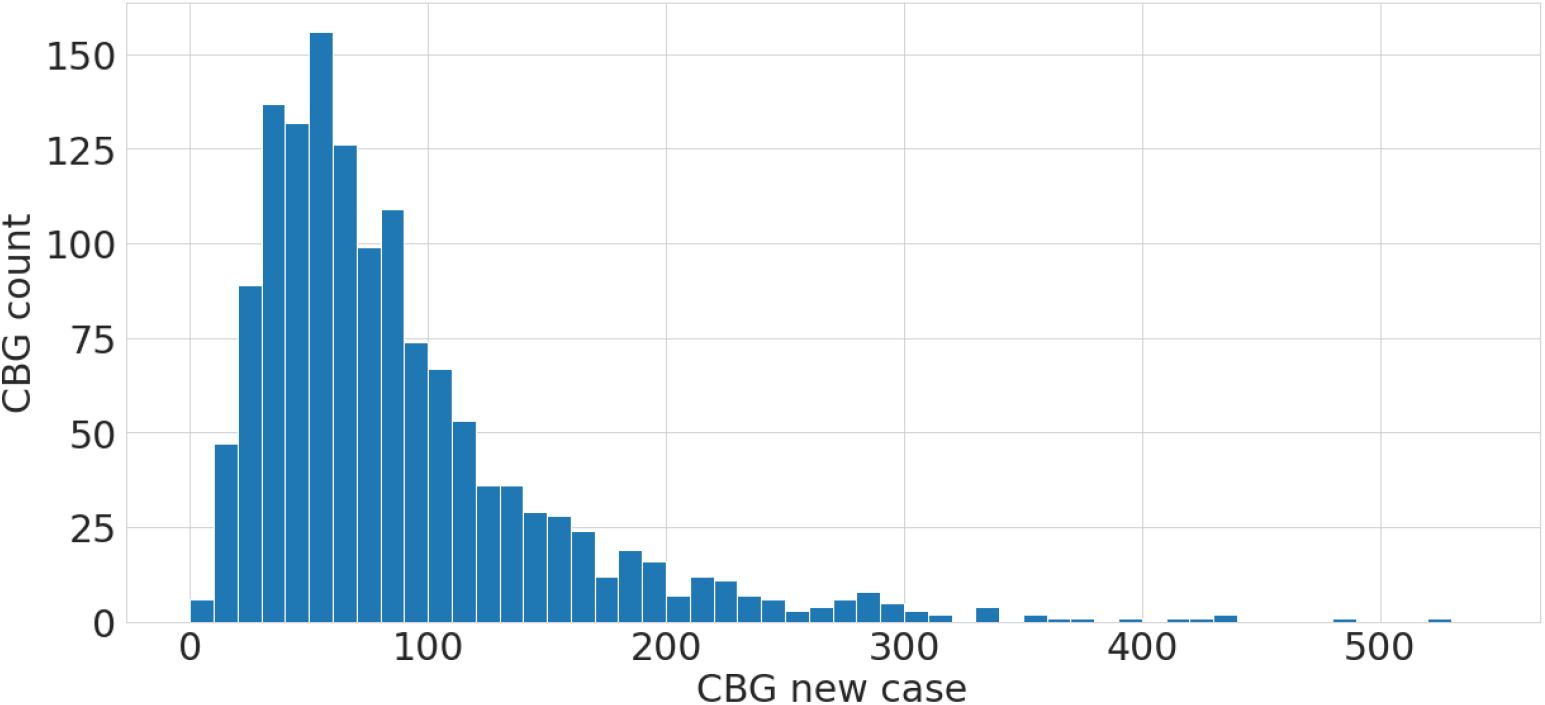
Histogram of CBG infection counts. Most CBGs have less than 100 COVID-19 infections that occurred in the simulation; a few CBGs have much higher infections. CBG: Census blockgroup.

**Figure A3.**
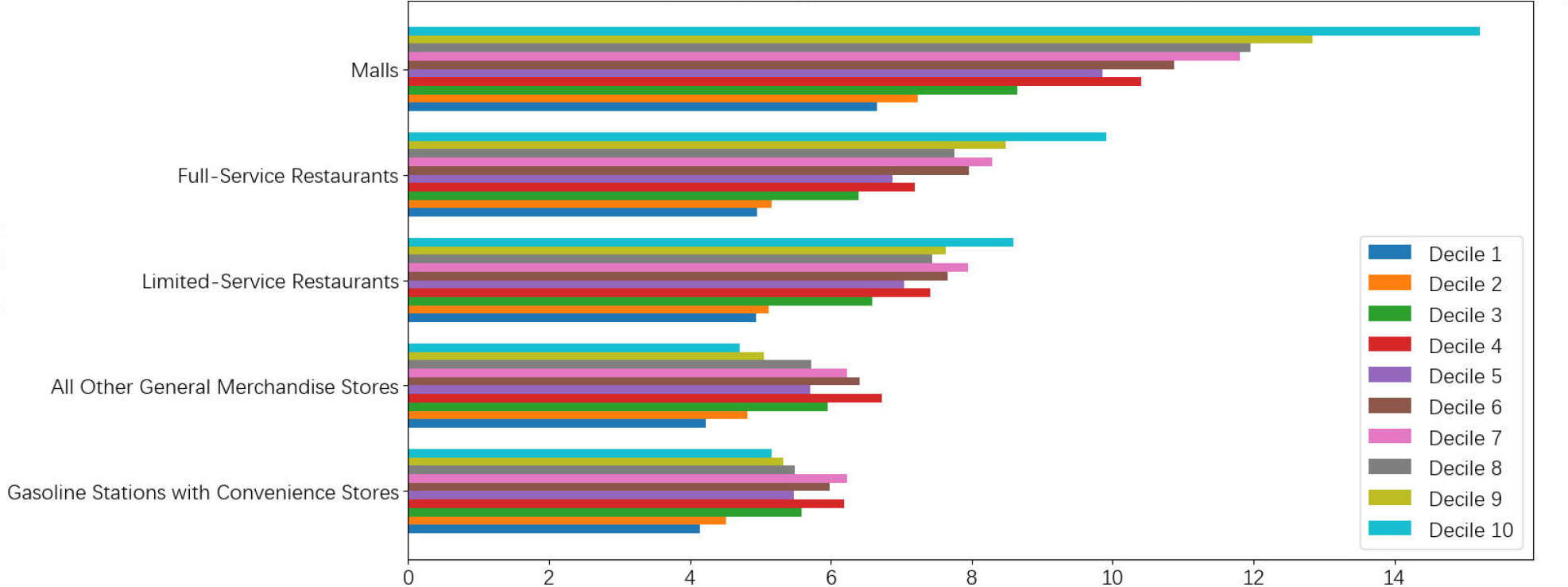
Per capita visits in the study area. The high-income household had more visits to malls and restaurants. (Decile 1: lowest median household income; Decile 10: highest median household income)

**Figure A4.**
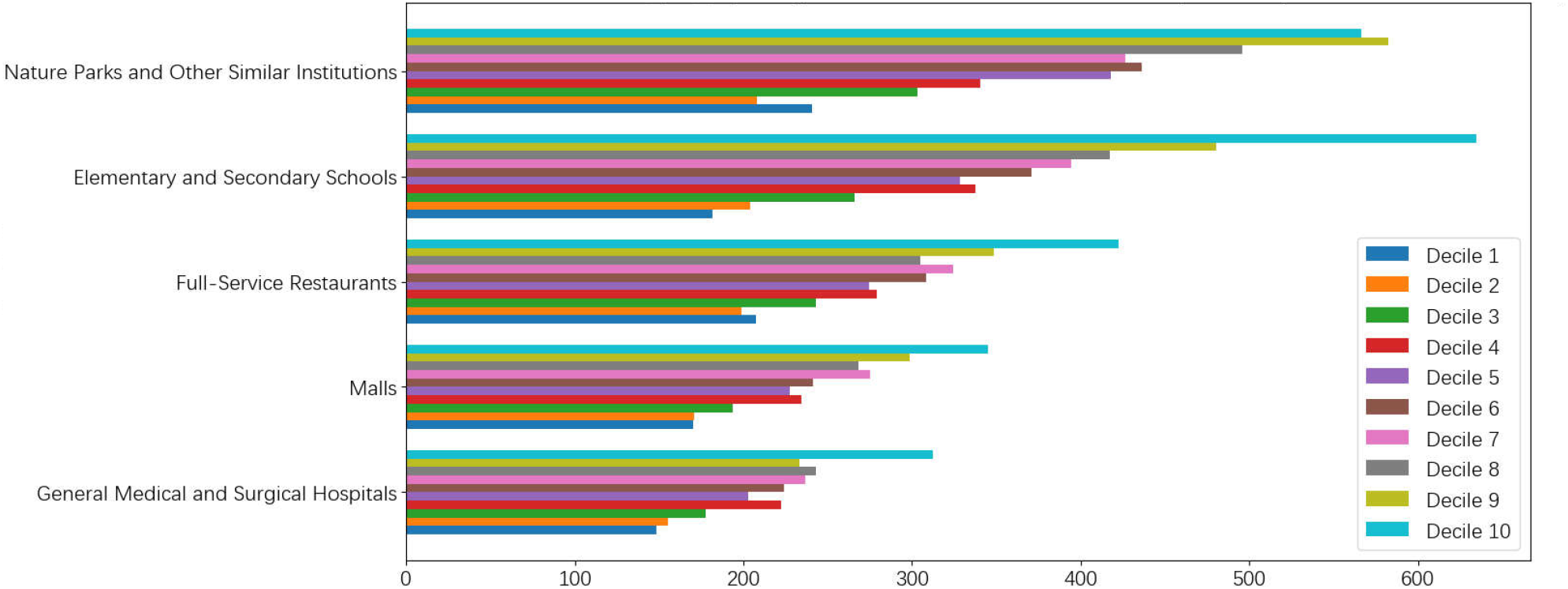
Per capita time spend (minute) in the study area. The high-income household spent more time on nature parks, schools, malls, and restaurants. (Decile 1: lowest median household income; Decile 10: highest median household income)

**Figure A5.**
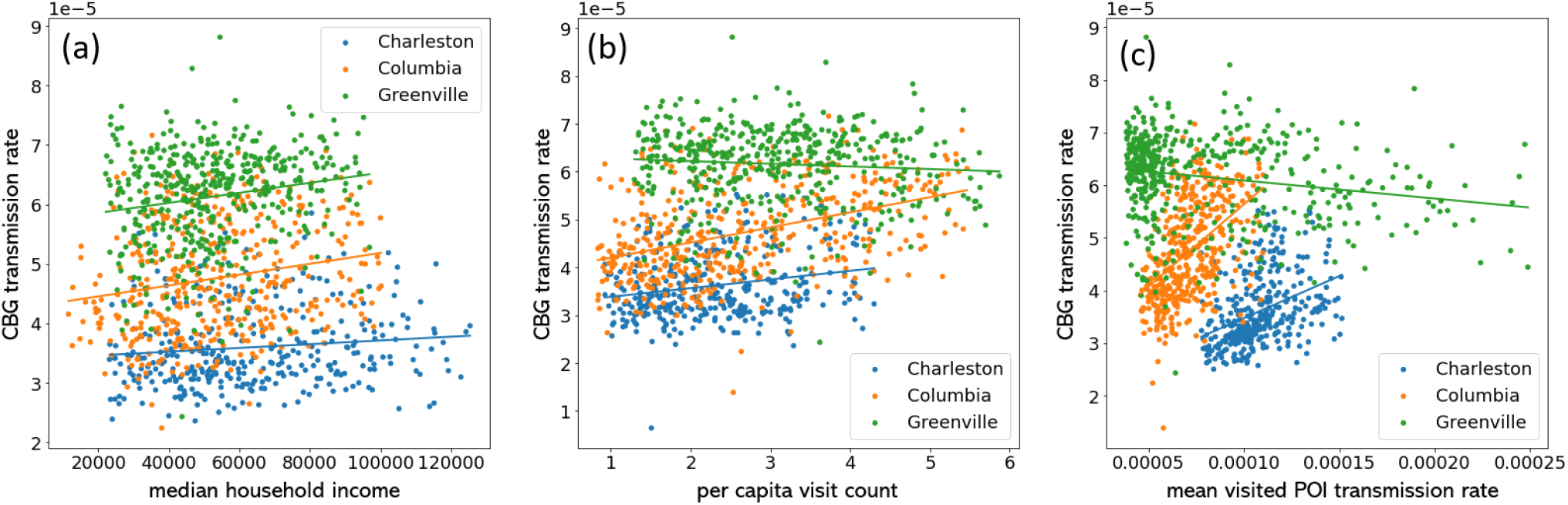
CBG transmission rates demonstrated different patterns among MSAs; the solid lines are the trend lines of the point cluster. (a) At the same CBG income level, Greenville had high transmission rates, and Charleston had low rates. (b) Charleston CBGs had fewer per capita visit counts and low transmission rates than Greenville and Columbia. (c) Greenville CBGs had high transmission rates, although their mean visited POI transmission rates were low; Charleston showed the opposite pattern. CBG: Census blockgroup; MSA: Metropolitan statistical areas.

**Table A2.**
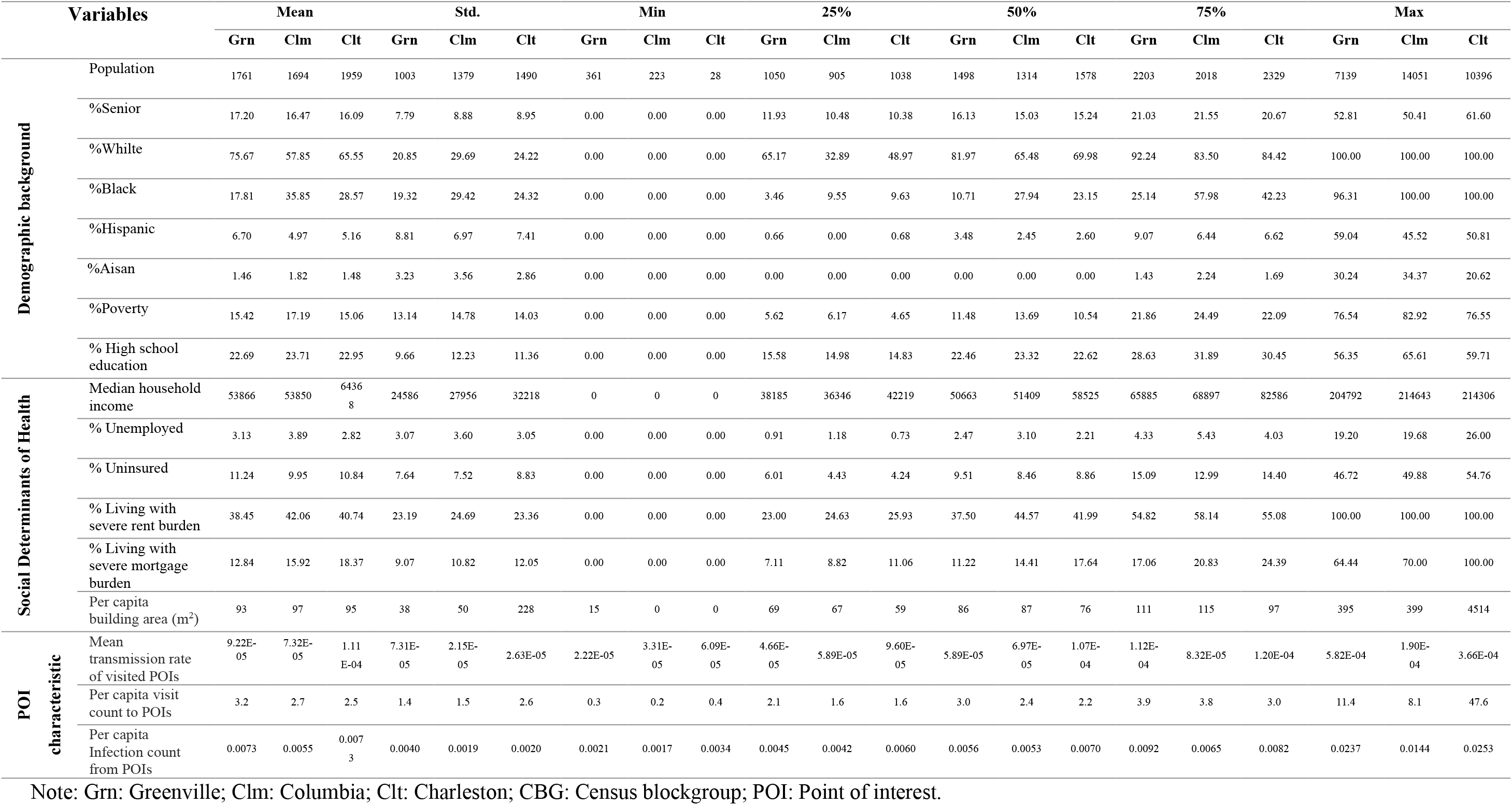
Descriptive statistics of the variables for correlation analysis of CBG transmission rate

|  | Variables | Mean |  |  | Std. |  |  | Min |  |  | 25% |  |  | 50% |  |  | 75% |  |  | Max |  |  |
| --- | --- | --- | --- | --- | --- | --- | --- | --- | --- | --- | --- | --- | --- | --- | --- | --- | --- | --- | --- | --- | --- | --- |
|  |  | Grn | Clm | Clt | Grn | Clm | Clt | Grn | Clm | Clt | Grn | Clm | Clt | Grn | Clm | Clt | Grn | Clm | Clt | Grn | Clm | Clt |
| Demographic background | Population | 1761 | 1694 | 1959 | 1003 | 1379 | 1490 | 361 | 223 | 28 | 1050 | 905 | 1038 | 1498 | 1314 | 1578 | 2203 | 2018 | 2329 | 7139 | 14051 | 10396 |
|  | %Senior | 17.20 | 16.47 | 16.09 | 7.79 | 8.88 | 8.95 | 0.00 | 0.00 | 0.00 | 11.93 | 10.48 | 10.38 | 16.13 | 15.03 | 15.24 | 21.03 | 21.55 | 20.67 | 52.81 | 50.41 | 61.60 |
|  | %White | 75.67 | 57.85 | 65.55 | 20.85 | 29.69 | 24.22 | 0.00 | 0.00 | 0.00 | 65.17 | 32.89 | 48.97 | 81.97 | 65.48 | 69.98 | 92.24 | 83.50 | 84.42 | 100.00 | 100.00 | 100.00 |
|  | %Black | 17.81 | 35.85 | 28.57 | 19.32 | 29.42 | 24.32 | 0.00 | 0.00 | 0.00 | 3.46 | 9.55 | 9.63 | 10.71 | 27.94 | 23.15 | 25.14 | 57.98 | 42.23 | 96.31 | 100.00 | 100.00 |
|  | %Hispanic | 6.70 | 4.97 | 5.16 | 8.81 | 6.97 | 7.41 | 0.00 | 0.00 | 0.00 | 0.66 | 0.00 | 0.68 | 3.48 | 2.45 | 2.60 | 9.07 | 6.44 | 6.62 | 59.04 | 45.52 | 50.81 |
|  | %Aisan | 1.46 | 1.82 | 1.48 | 3.23 | 3.56 | 2.86 | 0.00 | 0.00 | 0.00 | 0.00 | 0.00 | 0.00 | 0.00 | 0.00 | 0.00 | 1.43 | 2.24 | 1.69 | 30.24 | 34.37 | 20.62 |
|  | %Poverty | 15.42 | 17.19 | 15.06 | 13.14 | 14.78 | 14.03 | 0.00 | 0.00 | 0.00 | 5.62 | 6.17 | 4.65 | 11.48 | 13.69 | 10.54 | 21.86 | 24.49 | 22.09 | 76.54 | 82.92 | 76.55 |
|  | % High school education | 22.69 | 23.71 | 22.95 | 9.66 | 12.23 | 11.36 | 0.00 | 0.00 | 0.00 | 15.58 | 14.98 | 14.83 | 22.46 | 23.32 | 22.62 | 28.63 | 31.89 | 30.45 | 56.35 | 65.61 | 59.71 |
| Social Determinants of Health | Median household income | 53866 | 53850 | 64368 | 24586 | 27956 | 32218 | 0 | 0 | 0 | 38185 | 36346 | 42219 | 50663 | 51409 | 58525 | 65885 | 68897 | 82586 | 204792 | 214643 | 214306 |
|  | % Unemployed | 3.13 | 3.89 | 2.82 | 3.07 | 3.60 | 3.05 | 0.00 | 0.00 | 0.00 | 0.91 | 1.18 | 0.73 | 2.47 | 3.10 | 2.21 | 4.33 | 5.43 | 4.03 | 19.20 | 19.68 | 26.00 |
|  | % Uninsured | 11.24 | 9.95 | 10.84 | 7.64 | 7.52 | 8.83 | 0.00 | 0.00 | 0.00 | 6.01 | 4.43 | 4.24 | 9.51 | 8.46 | 8.86 | 15.09 | 12.99 | 14.40 | 46.72 | 49.88 | 54.76 |
|  | % Living with severe rent burden | 38.45 | 42.06 | 40.74 | 23.19 | 24.69 | 23.36 | 0.00 | 0.00 | 0.00 | 23.00 | 24.63 | 25.93 | 37.50 | 44.57 | 41.99 | 54.82 | 58.14 | 55.08 | 100.00 | 100.00 | 100.00 |
|  | % Living with severe mortgage burden | 12.84 | 15.92 | 18.37 | 9.07 | 10.82 | 12.05 | 0.00 | 0.00 | 0.00 | 7.11 | 8.82 | 11.06 | 11.22 | 14.41 | 17.64 | 17.06 | 20.83 | 24.39 | 64.44 | 70.00 | 100.00 |
|  | Per capita building area (m<sup>2</sup>) | 93 | 97 | 95 | 38 | 50 | 228 | 15 | 0 | 0 | 69 | 67 | 59 | 86 | 87 | 76 | 111 | 115 | 97 | 395 | 399 | 4514 |
| POI characteristic | Mean transmission rate of visited POIs | 9.22E-05 | 7.32E-05 | 1.11E-04 | 7.31E-05 | 2.15E-05 | 2.63E-05 | 2.22E-05 | 3.31E-05 | 6.09E-05 | 4.66E-05 | 5.89E-05 | 9.60E-05 | 5.89E-05 | 6.97E-05 | 1.07E-04 | 1.12E-04 | 8.32E-05 | 1.20E-04 | 5.82E-04 | 1.90E-04 | 3.66E-04 |
|  | Per capita visit count to POIs | 3.2 | 2.7 | 2.5 | 1.4 | 1.5 | 2.6 | 0.3 | 0.2 | 0.4 | 2.1 | 1.6 | 1.6 | 3.0 | 2.4 | 2.2 | 3.9 | 3.8 | 3.0 | 11.4 | 8.1 | 47.6 |
|  | Per capita Infection count from POIs | 0.0073 | 0.0055 | 0.0073 | 0.0040 | 0.0019 | 0.0020 | 0.0021 | 0.0017 | 0.0034 | 0.0045 | 0.0042 | 0.0060 | 0.0056 | 0.0053 | 0.0070 | 0.0092 | 0.0065 | 0.0082 | 0.0237 | 0.0144 | 0.0253 |

Note: Grn: Greenville; Clm: Columbia; Clt: Charleston; CBG: Census blockgroup; POI: Point of interest.

**Table A3.** Descriptive statistics of the variables for correlation analysis of POI transmission rate

| Variables | Mean |  |  | Std. |  |  | Min |  |  | 25% |  |  | 50% |  |  | 75% |  |  | Max |  |  |
| --- | --- | --- | --- | --- | --- | --- | --- | --- | --- | --- | --- | --- | --- | --- | --- | --- | --- | --- | --- | --- | --- |
|  | Grn | Clm | Clt | Grn | Clm | Clt | Grn | Clm | Clt | Grn | Clm | Clt | Grn | Clm | Clt | Grn | Clm | Clt | Grn | Clm | Clt |
| **POI area** | 569 | 1740 | 967 | 4300 | 101015 | 18389 | 3 | 1 | 0 | 27 | 26 | 24 | 52 | 54 | 52 | 122 | 136 | 122 | 124872 | 8167508 | 1256654 |
| **Total POI visits** | 636 | 630 | 549 | 1762 | 1688 | 1552 | 11 | 12 | 15 | 111 | 113 | 108 | 251 | 248 | 230 | 578 | 596 | 510 | 56115 | 55396 | 51734 |
| **Infection count from POIs** | 1.2 | 1.7 | 2.1 | 9.5 | 7.4 | 5.8 | 0.0 | 0.0 | 0.0 | 0.0 | 0.1 | 0.1 | 0.2 | 0.3 | 0.4 | 0.7 | 1.0 | 1.7 | 525.8 | 306.4 | 132.5 |

Note: Grn: Greenville; Clm: Columbia; Clt: Charleston; POI: Point of interest.

